# A novel and efficient machine learning Mendelian randomization estimator applied to predict the safety and efficacy of sclerostin inhibition

**DOI:** 10.1101/2024.01.30.24302021

**Authors:** Marc-André Legault, Jason Hartford, Benoît J. Arsenault, Archer Y. Yang, Joelle Pineau

## Abstract

Mendelian Randomization (MR) enables estimation of causal effects while controlling for unmeasured confounding factors. However, traditional MR’s reliance on strong parametric assumptions can introduce bias if these are violated. We introduce a new machine learning MR estimator named Quantile Instrumental Variable (IV) that achieves low estimation error in a wide range of plausible MR scenarios. Quantile IV is distinctive in its ability to estimate nonlinear and heterogeneous causal effects and offers a flexible approach for subgroup analysis. Applying Quantile IV, we investigate the impact of circulating sclerostin levels on heel bone mineral density, osteoporosis, and cardiovascular outcomes in the UK Biobank. Employing various MR estimators and colocalization techniques that allow multiple causal variants, our analysis reveals that a genetically predicted reduction in sclerostin levels significantly increases heel bone mineral density and reduces the risk of osteoporosis, while showing no discernible effect on ischemic cardiovascular diseases. Quantile IV contributes to the advancement of MR methodology, and the case study on the impact of circulating sclerostin modulation contributes to our understanding of the on-target effects of sclerostin inhibition.

## 2 Introduction

Instrumental Variable (IV) estimation is a technique used to estimate the causal effect of an exposure on an outcome of interest from observational data. While IV estimation relies on the strong (and untestable) assumption of access to a valid *instrumental variable*—some variable which is assumed to only affect the outcome of interest via the exposure—it is a powerful technique because, unlike most other causal inference strategies, it allows estimation even in the presence of unobserved confounders of the exposure–outcome relationship. Because IV estimation relies on different assumptions than other study designs, it can be used when other designs are not applicable or susceptible to bias. Studies relying on IVs are increasingly being be used to provide robust evidence when combined with designs that rely on different assumptions, to *triangulate* a causal effect from multiple sources [1].

Genetic variants can be used as IVs to infer causal effects in Mendelian randomization (MR) studies. MR leverages the fact that genetic variants are fixed throughout life and are unaffected by environmental factors that may have confounding effects. The use of genetic variants as IVs has enabled the estimation of the causal effect of lipoprotein fractions ([2, 3]), to predict the safety and efficacy of modulating drug targets ([4, 5]) or to estimate the causal effect of circulating proteins on diseases ([6, 7]). Despite these successes, MR studies may be biased either by failures of the untestable “exclusion restriction” assumption due to horizontal pleiotropy—when the genetic variant affects the outcome both via the exposure *and* some other pathway—or due to inappropriate assumptions on the functional form of the exposure–outcome relationship. Most recent work on MR has focused on the former, for example by allowing a fraction of the IVs to be invalid (*e*.*g*. [8–10]). However, few studies addressed the validity of the parametric assumptions made by MR estimators.

Most of the current MR estimators assume that both the genetic effect on the exposure and the causal effect of the exposure on the outcome are linear. Recent efforts have substantially relaxed these linearity assumptions, either by considering polynomial functional forms [11], locally linear effects [12, 13] or semi-parametric models [14]. These innovations are important because they allow non-constant treatment effects to be estimated. They do not assume that the effect of a unit increase in the exposure is necessarily constant over the full range of the exposure allowing for more complex but plausible dynamics such as threshold effects, diminishing returns or exponential effects. Despite the progress made in the field of nonlinear MR, current models still rely on strong assumptions including the assumption of a constant genetic effect amongst levels of the exposure and covariates [15–17]. Nonparametric IV estimators [18, 19] do not require restricting the functional form relating the exposure and the outcome beyond an additive assumption on the confounding, and as a result, they allow for effect heterogeneity between the IV and exposure, and between the exposure and outcome. Modeling effect heterogeneity allows estimating causal effects for specified subgroups of individuals from a single model fit by conditioning the estimate on the levels of other covariates. Estimation of conditional treatment effects is a powerful tool to anticipate effect heterogeneity, for example, by investigating sex differences, age differences or other clinically meaningful subgroup effects.

The development of nonparametric IV estimators has evolved independently from MR in the econometrics, statistics and machine learning literature and there has been limited efforts to bridge these worlds. Here, we harness recent developments in machine learning and nonparametric IV estimation to propose a new estimator named Quantile IV that performs well in the context of MR [18, 20]. Quantile IV, drawing from Hartford *et al*.’s DeepIV model, incorporates a crucial simplification for enhanced performance in MR contexts without any added statistical assumption. Specifically, DeepIV is a two-stage procedure that first models the conditional distribution of the exposure given then IVs and covariates. This involves fitting a probabilistic model, often parametrized by a neural network, and to sample values during training. In Quantile IV, we replace this step by a neural network quantile regression and use simple averaging of the predicted conditional quantiles to avoid sampling. In a realistic MR simulation setup, we show that our method outperforms the default DeepIV estimator and other instantiations of this approach [14]. Using simulated data, we show that our estimator achieves low error in all of the considered MR scenarios and we quantify the coverage and type 1 error rate of confidence intervals obtained by bootstrap aggregation. To evaluate our MR estimator in a real-world scenario, we evaluated the causal effect of a decrease in circulating sclerostin on bone and cardiovascular diseases using a two-sample approach within the UK Biobank. Sclerostin is the drug target of romosozumab, an anti-sclerostin monoclonal antibody used to prevent fractures in individuals with osteoporosis [21, 22]. Our investigation of possible cardiovascular adverse events aims to clarify safety concerns related with sclerostin inhibition stemming from the observation of a higher number of adjudicated serious cardiovascular events in the treatment arm of clinical trials of romosozumab [21–24].

## 3 Results

### 3.1 Evaluation of nonparametric IV estimators in realistic MR simulation scenarios

To evaluate the use of neural network-based nonparametric IV estimators in MR, we evaluated the performance of two recently proposed estimators (DeepIV [20] and DeLIVR [14]), and our new proposed method (Quantile IV); we also compared all three with the traditional linear two-stage least squares estimator. There is no single parameter describing the shape of the causal relationship between the exposure and outcome in nonlinear settings. We use the square root of the mean squared error (RMSE) between the true causal relationship and the IV regression function to evaluate the performance of the different estimators. The RMSE is taken over evenly spaced points spanning the range of the exposure. We computed this metric for 200 simulation replicates, and we consider scenarios varying the causal relationship shape, the sample size, the variance explained in the exposure by the IVs, the strength of confounding and the number of IVs (Table 1). We report the results over the range of the exposure encompassing 95% of the distribution (Figure 1) but observed similar results over the full range (Supplementary Figure A1). Quantile IV was competitive and achieved low RMSE across all of the simulation scenarios. The linear estimate (two-stage least squares, 2SLS) is provided as a baseline comparison that is not expected to outperform the nonparametric estimators except in the linear model. Furthermore, we observed that the DeepIV estimator had high error and variability across the considered parameters prompting us to focus on DeLIVR and Quantile IV for quantitative comparisons. We used t-tests paired by simulation replicates to compare DeLIVR and Quantile IV (Table 2 and Supplementary Table A1). In most of our simulation scenarios, Quantile IV significantly outperformed DeLIVR at the nominal P-value threshold of 0.01 (12/15 scenarios when considering 95% of the exposure range and 9/15 scenarios when considering the full range). DeLIVR significantly outperformed Quantile IV when the number of instruments was set to 100, the largest number of IVs considered in our study. In all our simulations, there was a linear and homogeneous effect of the IVs on the exposure, which is an assumption made by DeLIVR, favoring this model.

**Table 1:** Values taken by the different parameters in the simulation study. We repeated every simulation 200 times and the bold values indicate the reference values for the parameters. The reference value represents the fixed value of a simulation parameter used when other simulation parameters are varied.

| Simulation parameter | Simulated Values |
| --- | --- |
| Structural equation | $0.4x$, **$0.2(x - 1)^2$**, $\max(x - 2, 0)$ |
| Sample size | 10 000, **50 000**, 100 000 |
| Heritability of the exposure | **0.1**, 0.2, 0.5 |
| Error correlation (confounding strength) | -0.6, **0.3**, 0.6 |
| Number of independent instrumental variables | 2, **10**, 100 |

**Table 2:** Comparison between the mean square root of the mean squared error between DeLIVR and Quantile IV across the different Mendelian randomization simulation scenarios and over the central 95% of the empirical exposure distribution. The P-value is from a paired t-test by simulation replicate.

| Simulation parameter | Sim. value | DeLIVR Mean RMSE (s.d.) | Quantile IV Mean RMSE (s.d.) | P-value |
| --- | --- | --- | --- | --- |
| Sample size | 10,000 | 0.21 (0.06) | 0.22 (0.10) | 0.094 |
| | 50,000 | 0.17 (0.03) | **0.13 (0.04)** | $2.81 \times 10^{-22}$ |
| | 100,000 | 0.18 (0.04) | **0.11 (0.02)** | $1.34 \times 10^{-57}$ |
| Instrument strength | 0.05 | 0.18 (0.04) | **0.15 (0.05)** | $1.50 \times 10^{-10}$ |
| | 0.1 | 0.17 (0.03) | **0.13 (0.04)** | $2.74 \times 10^{-24}$ |
| | 0.5 | 0.21 (0.04) | **0.12 (0.02)** | $3.01 \times 10^{-71}$ |
| Causal relationship shape | Linear | 0.08 (0.04) | **0.06 (0.03)** | $1.79 \times 10^{-5}$ |
| | Quadratic | 0.18 (0.04) | **0.12 (0.03)** | $7.89 \times 10^{-38}$ |
|  | Threshold | 0.06 (0.03) | 0.06 (0.03) | 0.061 |
| Confounding | -0.6 | 0.18 (0.03) | **0.15 (0.05)** | $8.53 \times 10^{-12}$ |
| | 0.3 | 0.18 (0.09) | **0.13 (0.04)** | $4.07 \times 10^{-15}$ |
| | 0.6 | 0.18 (0.03) | **0.11 (0.03)** | $9.62 \times 10^{-58}$ |
| Number of instruments | 2 | 0.23 (0.09) | **0.17 (0.09)** | $3.38 \times 10^{-20}$ |
| | 10 | 0.17 (0.02) | **0.12 (0.04)** | $5.58 \times 10^{-40}$ |
| | 100 | **0.16 (0.01)** | 0.22 (0.04) | $2.35 \times 10^{-41}$ |

**Figure 1:**
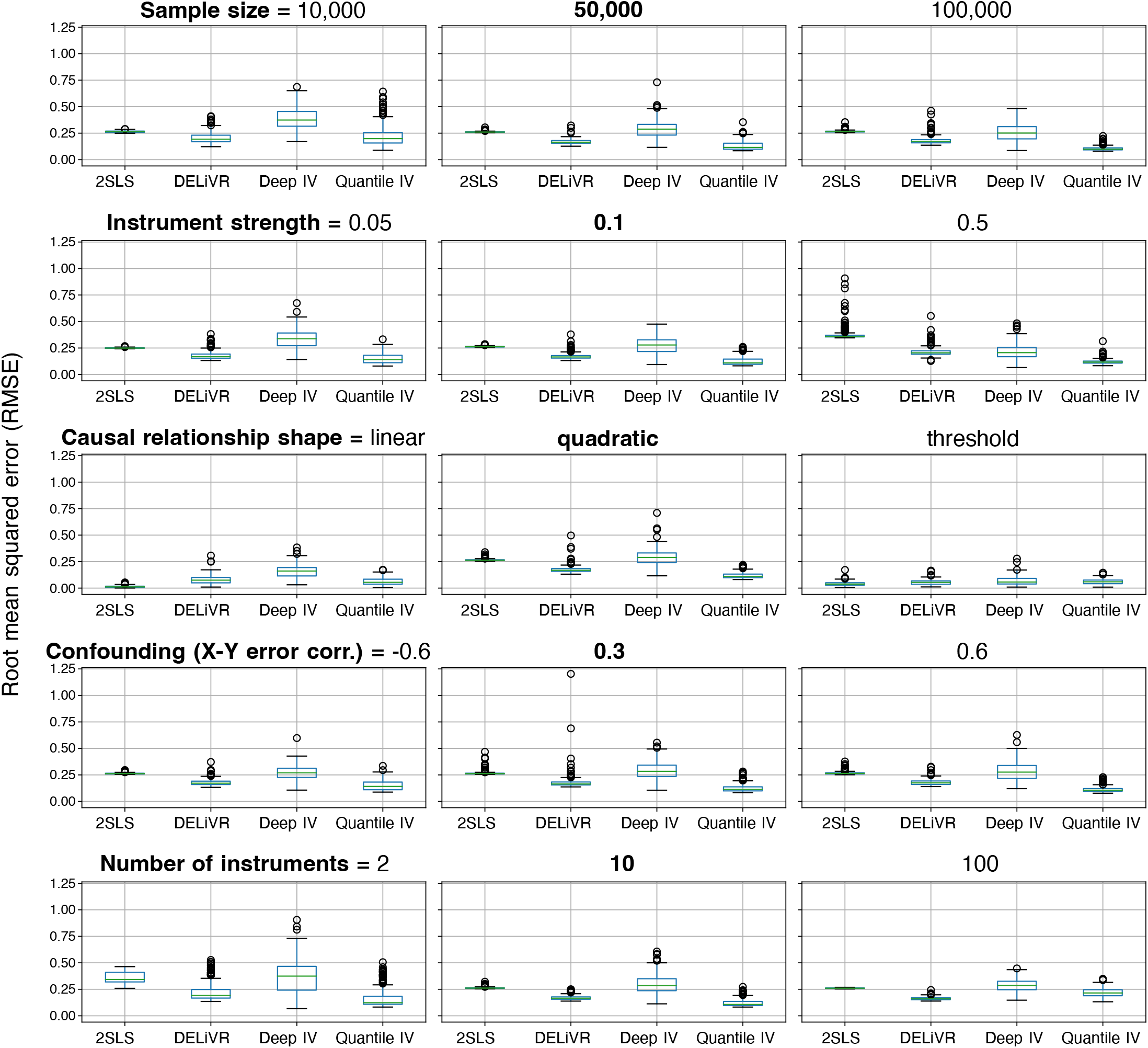
Root mean squared error between the estimated IV regression and the true causal function over a grid spanning 95% of the empirical range of the exposure. The boxplot for every estimator represents variability over 200 simulation replicates. The simulation parameter values in bold correspond to the reference values. 2SLS: Two-stage least squares, DeLIVR [14], DeepIV [20], Quantile IV: proposed method.

A drawback of many nonparametric IV methods is that they do not easily allow for the computation of confidence intervals or provide means to quantify uncertainty. We evaluated the use of bootstrap aggregation (bagging) to construct confidence intervals focusing on the simulation scenario consisting of the baseline value for all of the simulation parameters (Table 1). We observed that coverage of the average treatment effect (ATE) for a unit increment in the exposure was close or upwards of the nominal value across most of the exposure range (Supplementary Figure A2). We recommend reporting effects within the central range of the exposure, where coverage was more consistent (*e*.*g*. between the 2.5th and 97.5th percentiles of the empirical distribution of the exposure). We also observed a surprising drop in coverage below the nominal level for exposure values near 0 despite a low absolute error in the estimation of the average treatment effect (Supplementary Figure A2). We attribute this to overconfidence of the bagging confidence interval in this region which could be due to factors related to the selection of the neural network parameter initialization or architecture. We used a similar strategy to evaluate the false positive rate and observed that our estimator did not exceed the nominal level (Supplementary Figure A3). Across the range of the exposure, we observed an increase of the type 1 error rate close to the nominal level at a similar location to the decrease in coverage. This observation is concordant with the hypothesis that the neural network parameter initialization or architecture may be further optimized to improve on these finite sample results.

### 3.2 Mendelian randomization study population

We used a two-sample design within the UK Biobank (split sample) for this study. We used a subset of 42,830 participants with available sclerostin measurements to estimate all the effect related to this measurement. We used a non-overlapping subset of 370,218 participants to estimate all the genetic effects related to our outcomes of interest. Descriptive statistics for both datasets are shown in Supplementary Table A2.

### 3.3 Genetic association and colocalization analysis of circulating sclerostin and outcomes in the UK Biobank

To identify *cis*-pQTLs associated with circulating sclerostin levels, we conducted a genetic association analysis of 1,449 common variants at the *SOST* locus in the UK Biobank (Supplementary Figure A4). Our goal was to select strong genetic predictors of sclerostin levels for subsequent MR analyses by using finemapping to select IVs. A SuSiE analysis [25] revealed two 95% credible sets with log_10_ of the Bayes factor of 10.3 and 5.0. The first set contained 17 variants and the variant with the largest posterior inclusion probability (PIP) was rs6416905 (PIP = 0.078). The second set contained 6 variants and rs66838809 had the largest posterior inclusion probability (PIP = 0.48). Variants within the two credible sets were in high linkage disequilibrium (LD) with a mean *r*^2^ = 1 for the first set and mean *r*^2^ = 0.89 for the second set. We selected the two variants with the highest PIP for subsequent MR analyses as they were the best representatives of the two independent signals identified by SuSiE. As a sensitivity analysis, we also used a forward stepwise regression procedure and obtained concordant results (Supplementary Note). The estimated effects of the two selected IVs on circulating sclerostin levels are presented in Supplementary Table A3.

We used the same genetic association and fine-mapping procedure for the outcomes of interest, namely heel bone mineral density, osteoporosis, and cardiovascular outcomes (PCI/CABG, MI, ischemic stroke and acute CAD). The association P-values expressed with respect to the sclerostin decreasing allele are shown in Figure 2 along with measured LD in the region. We identified significant associations with heel bone mineral density and the top association was with rs66838809 (chr17:41,798,621 G/A), whose “A” allele had 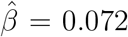, 95% CI (0.061, 0.083), *P* = 5.8 *×*10^−38^ (Table 3). This variant is also a lead variant for one of the sclerostin pQTL credible sets (Supplementary Table A3). We identified an association between genetic variants at the *SOST* locus and osteoporosis and the lead variant was again rs66838809 (chr17:41,798,621 G/A) whose “A” allele had OR = 0.83, 95% CI (0.80, 0.87), *P* = 3.8*×*10^−17^. To formally test whether the observed associations were due to shared causal variants with the sclerostin levels, we conducted a colocalization analysis [26]. The colocalization algorithm estimates the posterior probability (PP) of four scenarios denoted by H_1_ to H_4_ including the existence of a shared causal variants underlying the association signal in both of the considered traits (H_4_). It is possible to test for these hypotheses assuming a single causal variant, or to use finemapping and test pairs of credible sets to allow for multiple causal variants per region [26]. Colocalization analysis revealed strong evidence of a shared causal variant between heel bone mineral density and circulating sclerostin levels. The two sclerostin pQTL credible sets colocalized with heel bone mineral density credible set with PPH_4_ = 0.964 and 0.997. There was also strong evidence for colocalization between osteoporosis and one of the pQTL credible sets with posterior probabilities of a shared causal variant of 0.996. The second sclerostin pQTL credible set (represented by rs6416905) had no evidence of colocalization with osteoporosis (PPH_4_ = 0.013). Overall, there was robust evidence of colocalization between genetic associations with circulating sclerostin levels, heel bone mineral density and osteoporosis recapitulating the known protective effect of pharmacological sclerostin inhibition in osteoporosis [21, 22].

**Table 3:** Association statistics between the two lead variants for the sclerostin pQTL credible sets identified by SuSiE and quantitative traits and diseases. All of the effects are presented with respect to the sclerostin reducing allele. The UK Biobank associations were estimate in the current study and we present associations published by the GTEx (sclerostin expression) and CARDIoGRAMplusC4D (cardiovascular diseases) consortia.

| Dataset | rs6416905 |  | rs66838809 |  |
| --- | --- | --- | --- | --- |
|  | Coef. / OR (95% CI) | P-value | Coef. / OR (95% CI) | P-value |
| *UK Biobank quantitative traits* |  |  |  |  |
| pQTL | -0.059 (-0.072, -0.046) | $1.50 \times 10^{-18}$ | -0.097 (-0.120, -0.074) | $1.80 \times 10^{-16}$ |
| Heel bone mineral density | 0.032 (0.026, 0.039) | $3.30 \times 10^{-25}$ | 0.072 (0.061, 0.083) | $5.80 \times 10^{-38}$ |
| *UK Biobank diseases (OR scale)* |  |  |  |  |
| Osteoporosis | 0.954 (0.934, 0.976) | $3.10 \times 10^{-5}$ | 0.834 (0.800, 0.870) | $3.80 \times 10^{-17}$ |
| Myocardial infarction | 0.988 (0.967, 1.010) | 0.28 | 1.017 (0.979, 1.057) | 0.39 |
| Acute CAD | 1.000 (0.981, 1.019) | 0.99 | 1.042 (1.008, 1.078) | 0.02 |
| Ischemic stroke | 0.986 (0.955, 1.018) | 0.38 | 1.002 (0.946, 1.061) | 0.95 |
| PCI/CABG | 1.007 (0.984, 1.030) | 0.58 | 1.064 (1.022, 1.108) | 0.003 |
| *Summary statistics (consortia) quantiative traits* |  |  |  |  |
| GTEx V8 eQTL (artery tibial) | -0.253 (-0.300, -0.206) | $6.00 \times 10^{-23}$ | -0.311 (-0.410, -0.211) | $2.40 \times 10^{-9}$ |
| GTEx V8 eQTL (artery aorta) | -0.247 (-0.319, -0.176) | $7.00 \times 10^{-11}$ | -0.401 (-0.542, -0.261) | $5.00 \times 10^{-8}$ |
| *Summary statistics (consortia) diseases (OR scale)* |  |  |  |  |
| CARDIoGRAM MI | 1.002 (0.981, 1.024) | 0.83 | 1.043 (0.998, 1.091) | 0.06 |
| CARDIoGRAM CAD | 0.991 (0.973, 1.010) | 0.37 | 0.997 (0.958, 1.038) | 0.90 |
| CARDIoGRAM & UKB CAD | 0.994 (0.978, 1.011) | 0.48 | 0.999 (0.966, 1.033) | 0.96 |

**Figure 2:**
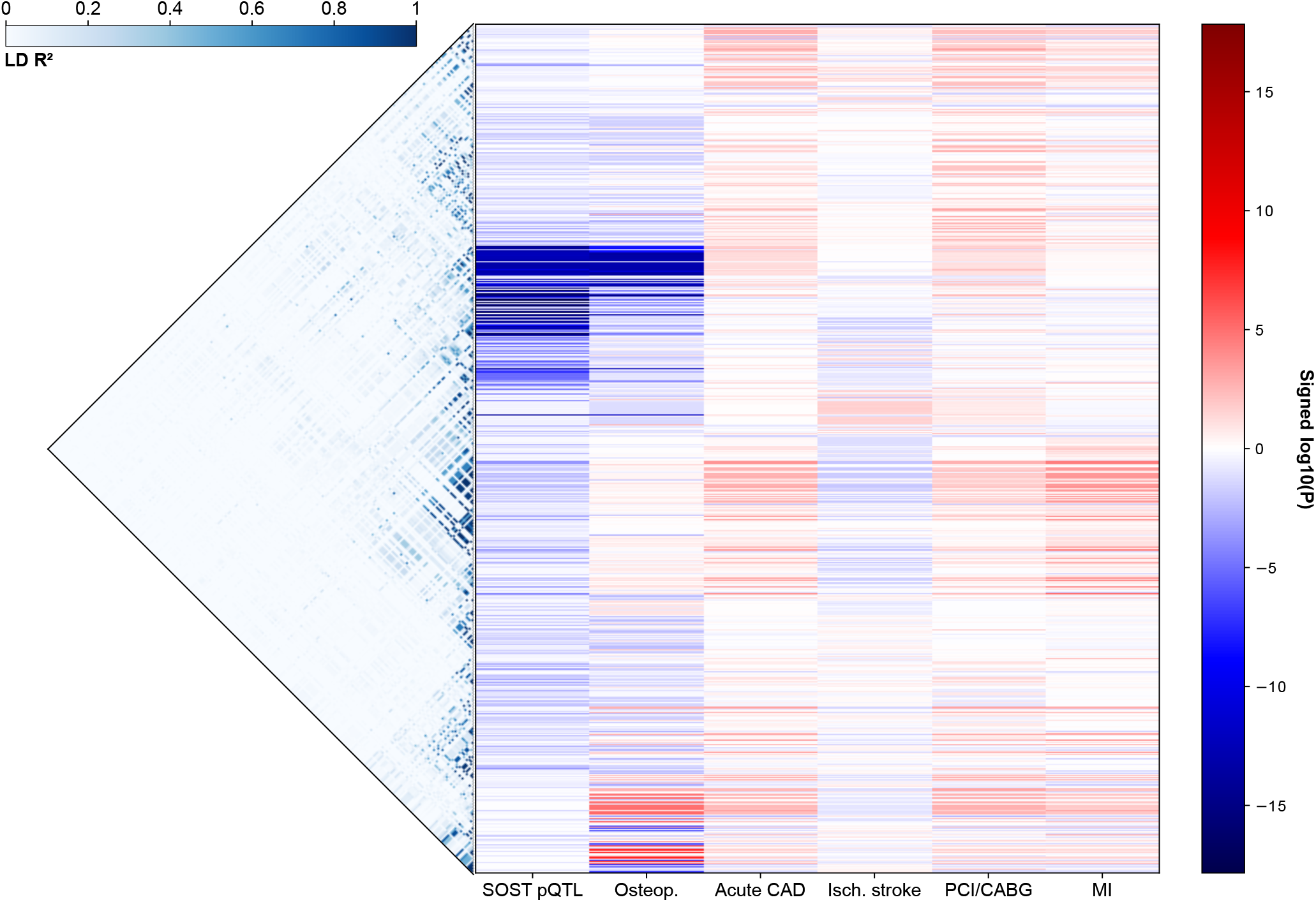
Association between common genetic variants at the *SOST* locus (chr17:41,631,099-42,236,156) and the exposure and outcomes considered in our MR study in the UK Biobank. The variants are ordered with respect to their genomic coordinate and the leftmost part of the plot shows the observed LD between the variants. The color lines represent association P-values for every genetic variant and phenotype colored with respect to the sign of the regression coefficient (red for trait increasing and blue for trait decreasing). The sclerostin decreasing allele is the coded allele throughout.

There was evidence of genetic associations at the *SOST* locus with some of the considered cardiovascular outcomes (Figure 2). The outcome with the most significant genetic association was myocardial infarction and the “T” allele of the lead variant, rs75086002 (chr17:42,021,918 C/T), had an OR of 0.92 95% CI (0.89, 0.96) *P* = 5.9 *×* 10^−6^. This association does not reach the genome-wide significance threshold, but it did cross the conservative Bonferroni threshold considering multiple testing of 1,449 variants (3.5 *×* 10^−5^ = 0.05/1449). Using SuSiE, we inferred a 90% credible set for this association signal, but it did not colocalize with the sclerostin pQTL credible sets (PPH_4_ with the rs6416905 credible set = 0.005 and PPH_4_ with the rs66838809 credible set = 0.004). Visual inspection of the association signals and inferred credible sets is also consistent with the hypothesis of distinct association signals for myocardial infarction and sclerostin levels (Supplementary Figure A5). The most probable hypothesis according to colocalization analysis is that the associations are due to distinct genetic variants (PPH_3_ = 0.78). Because inference of credible sets can be statistically challenging, we also tested colocalization assuming the single causal variant assumption. The results were concordant with the absence of colocalization between sclerostin pQTLs and cardiovascular diseases (largest PPH_4_ = 0.01 with ischemic stroke). We retrieved the summary association statistics between our two selected sclerostin pQTLs and the considered outcomes in the UK Biobank and in data from the CARDIoGRAMplusC4D consortium which includes over 70,000 cases of CAD (Table 3). In our UK Biobank analysis, there was a nominally significant association of rs66838809 with acute CAD (*P* = 0.02) and PCI/CABG (*P* = 0.003), but no evidence of association in data from the CARDIoGRAMplusC4D consortium (P-value for the largest CAD study was 0.48 for rs6416905 and 0.96 for rs66838809).

Previous studies have considered eQTLs of sclerostin levels as instruments in MR studies [23, 27]. We sought to evaluate whether there was a shared genetic basis to the regulation of sclerostin eQTLs and circulating sclerostin levels. The selected sclerostin pQTLs were robustly associated with *SOST* expression in aorta and tibial arteries (*P ≤* 5 *×* 10^−8^, Table 3). We observed that the eQTLs and pQTLs were located near each other on the chromosome and were significantly associated with both sclerostin gene expression and circulating levels (Supplementary Figures A6 to A8). Statistical colocalization analysis however suggests that the underlying causal variants are different (Max PPH_4_ = 0.12 for aorta and 0.004 for tibial artery, Supplementary Figure A9). This finding could be due to the presence of different genetic regulatory mechanisms behind sclerostin gene expression in comparison to circulating protein levels, but it could also be due to limited statistical power in GTEx or a mismatch between the GTEx and UK Biobank populations hampering statistical finemapping analyses.

We conclude that association analysis identified strong *cis*-pQTLs of circulating sclerostin levels that co-localize with genetic associations with heel bone mineral density and osteoporosis. Despite some evidence of genetic associations with cardiovascular outcomes at the *SOST* locus, evidence from large consortia of CAD and colocalization analyses suggest that they may be unrelated to genetic variants influencing the regulation of circulating sclerostin levels.

### 3.4 Mendelian randomization of the effect of circulating sclerostin on bone and cardiovascular diseases

To estimate the causal effect of a genetically predicted reduction in circulating sclerostin levels on bone and cardiovascular traits and outcomes, we considered three complementary approaches suitable for the *cis*-MR context [28]. Using the finemapped sclerostin pQTLs as IVs, we used the inverse variance weighted (IVW) estimator accounting for LD and our Quantile IV nonparametric estimator. As a complementary approach, we used PC-GMM, a novel estimator that can leverage all of the variants in the region [29]. All of the methods estimated that reducing circulating sclerostin would result in a statistically significant increase in heel bone mineral density and a reduction in the risk of osteoporosis (Table 4). However, the magnitude of the estimates was different across estimators. We report the estimated effect for both a 1 s.d. reduction in sclerostin levels about the mean, and a 2 s.d. reduction about the mean (Table 4). These two contrasts correspond, assuming normality in the distribution of circulating sclerostin levels, to a reduction from the mean to the level of the bottom 16% or the bottom 2% of the distribution of the exposure. The IVW estimator suggests that a 2 s.d. reduction in sclerostin levels about the mean reduces the odds of osteoporosis by a surprising 92% (OR = 0.085). For the same change in circulating sclerostin levels, Quantile IV estimates an OR = 0.626. Upon visual inspection of the Quantile IV estimate, there was no evidence of pronounced nonlinearity (Figure 3). When considering heel bone mineral density and osteoporosis as outcomes, Quantile IV estimated smaller, albeit significant, causal effects than conventional linear methods.

**Table 4:** Mendelian randomization estimate of a 1 s.d. or 2 s.d. reduction in sclerostin levels about the mean on heel bone mineral density and osteoporosis. Three different MR estimators are considered, and the estimates are from a two-sample MR within the UK Biobank.

| Phenotype | Estimator | ATE 1 s.d. reduction (95% CI) | ATE 2 s.d. reduction (95% CI) |
| --- | --- | --- | --- |
| Heel BMD | IVW | 0.639 (0.554, 0.723) | 1.277 (1.108, 1.446) |
|  | PC-GMM | 0.526 (0.162, 0.889) | 1.051 (0.324, 1.778) |
|  | Quantile IV | 0.122 (0.117, 0.125) | 0.236 (0.230, 0.243) |
| Osteoporosis (OR scale) | IVW | 0.291 (0.213, 0.398) | 0.085 (0.045, 0.158) |
|  | PC-GMM | 0.349 (0.182, 0.670) | 0.122 (0.033, 0.449) |
|  | Quantile IV | 0.794 (0.612, 0.949) | 0.626 (0.373, 0.857) |

**Figure 3:**
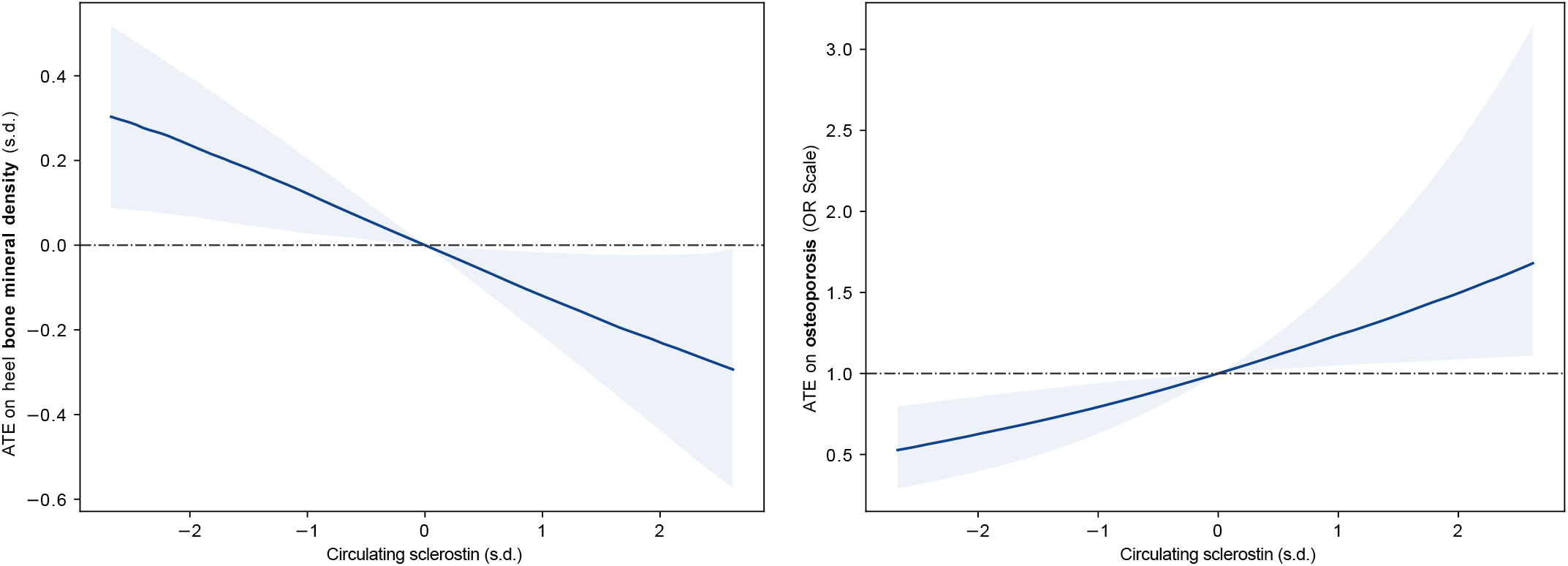
Quantile IV estimate of the average effect varying the levels of circulating sclerostin about the mean on heel bone mineral density and osteoporosis in the UK Biobank. The shaded region corresponds to 90% bootstrap confidence intervals. The plots cover the central 99% of the exposure range.

Nonparametric IV estimation enables the estimation of conditional average treatment effects (CATE) without explicitly specifying an interaction model. Such effects represent the average effect of a treatment in a specific subset of individuals and can be used to assess treatment response heterogeneity. To evaluate if the causal effect of varying circulating sclerostin levels differs across levels of covariables, we estimated CATEs in men, women and in individuals with different values of age at baseline. Age did not modify the effect of sclerostin inhibition on heel bone mineral density or osteoporosis (*P* = 0.43 and *P* = 0.35, respectively). However, we observed sex heterogeneity as the CATE for a 1 s.d. decrease in circulating sclerostin levels was 0.15 for women vs 0.09 for men (interaction *P* = 2.3 *×* 10^−22^, Supplementary Table A4, Supplementary Figure A10). We observed directionally concordant sex-differences on the effect of sclerostin inhibition on osteoporosis with interaction *P* = 0.04, but the effect difference was small.

When estimating the causal effect of a reduction of sclerostin levels on cardiovascular outcomes there was disagreement between the results obtained by different estimators. The IVW and PC-GMM estimated a nominally significant increase in the risk of PCI/CABG (*P*_IVW_ = 0.026, *P*_PC-GMM_ = 0.017, Supplementary Figures A11 and A12) and PC-GMM estimated a nominally significant increase in the risk of acute CAD (*P* = 0.021, Supplementary Figure A12). Quantile IV, on the other hand, estimated a nominally significant protective effect on PCI/CABG (OR = 0.74, 95% CI (0.52, 0.97), Supplementary Figures A13 and A14). However, we interpret these results with care as estimates from the three estimators were heterogeneous and there was no supporting colocalization evidence for shared causal variants between sclerostin pQTLs and cardiovascular diseases in our previous analysis. We attribute these results to bias due to LD with other genetic variants that have effects on the outcome independently from sclerostin levels and investigate this in the next section.

### 3.5 Mendelian randomization analyses accounting for pleiotropic effects

In the MR analyses adjusted for age, sex, and ancestry principal components, we observed conflicting effects for PCI/CABG despite limited evidence of a genetic associations between *SOST* variants and this outcome. The top association was with rs370088062, chr17:41,657,403 “CT” to “C” deletion, with *P* = 2.9 *×* 10^−4^. This variant showed no association with circulating levels of sclerostin (*P* = 0.40), indicating that the observed effects in Mendelian Randomization might be biased due to LD. More precisely, if the IVs are in LD with other genetic variants that have effects on the outcome independently from sclerostin levels, the *exclusion restriction* assumption will be violated biasing the MR estimates. This problem is particularly challenging in the *cis*-MR context because of LD and the limited number of candidate IVs. To identify the variants most at risk of violating the MR assumptions, we repeated the association analysis with PCI/CABG adjusting for all of the pQTLs identified in the stepwise conditional analysis (Supplementary Note). We then compared the association P-values before and after adjustment for the pQTL associations under the premise that variants whose association is unattenuated may influence PCI/CABG risk through pathways unrelated to sclerostin (Supplementary Figure A15). This analysis revealed a group of correlated variants including rs113533733 that had low association P-values with PCI/CABG after adjusting for the sclerostin pQTL variants.

We sought to confirm that rs113533733 may have effects on other genes by consulting the Open Targets Genetics platform [30]. This online resource includes a variant to gene prioritization module based in part on the distance to transcription start sites and pQTL, splice QTL (sQTL) and eQTL data. On this platform, the most likely gene assigned to rs113533733 is *MPP3* (score = 0.31) with support from sQTL and eQTL data. The other prioritized genes are *DUSP3* (score = 0.18), *CFAP97D1* (nearest gene, score = 0.15) and *MPP2* (score = 0.13). There is no evidence linking *SOST* to rs113533733 except for its distance to the transcription start site of 41 kb and the assigned score is 0.06. In GTEx V8, the strongest eQTL for this variant was with *MPP3* in the heart left ventricle tissue (*P* = 2.1 *×* 10^−5^). Considering this external evidence and our association analysis conditional on sclerostin pQTLs, we concluded that rs113533733 may induce bias in the MR analysis and repeated our MR estimation adjusting for this variant.

In the MR analysis adjusted for rs113533733, the effect of a 1 s.d. reduction in circulating sclerostin levels on heel bone mineral density and osteoporosis remained significant with no attenuation in the P-value for all methods (Figure 4 and Supplementary Table A5). After adjustment for rs113533733 the estimated causal effects of sclerostin levels on the considered cardiovascular diseases were null for PC-GMM and Quantile IV (Figure 4, Supplementary Figure A16). The IVW had an inconsistent estimate for the effect of sclerostin reduction on PCI/CABG (OR = 1.41, 95% CI (1.02, 1.96), *P* = 0.04), but the large confidence interval, discordance with the other estimators, absence of effect with related traits and lack of support from colocalization analyses suggest the true effect is likely null.

**Figure 4:**
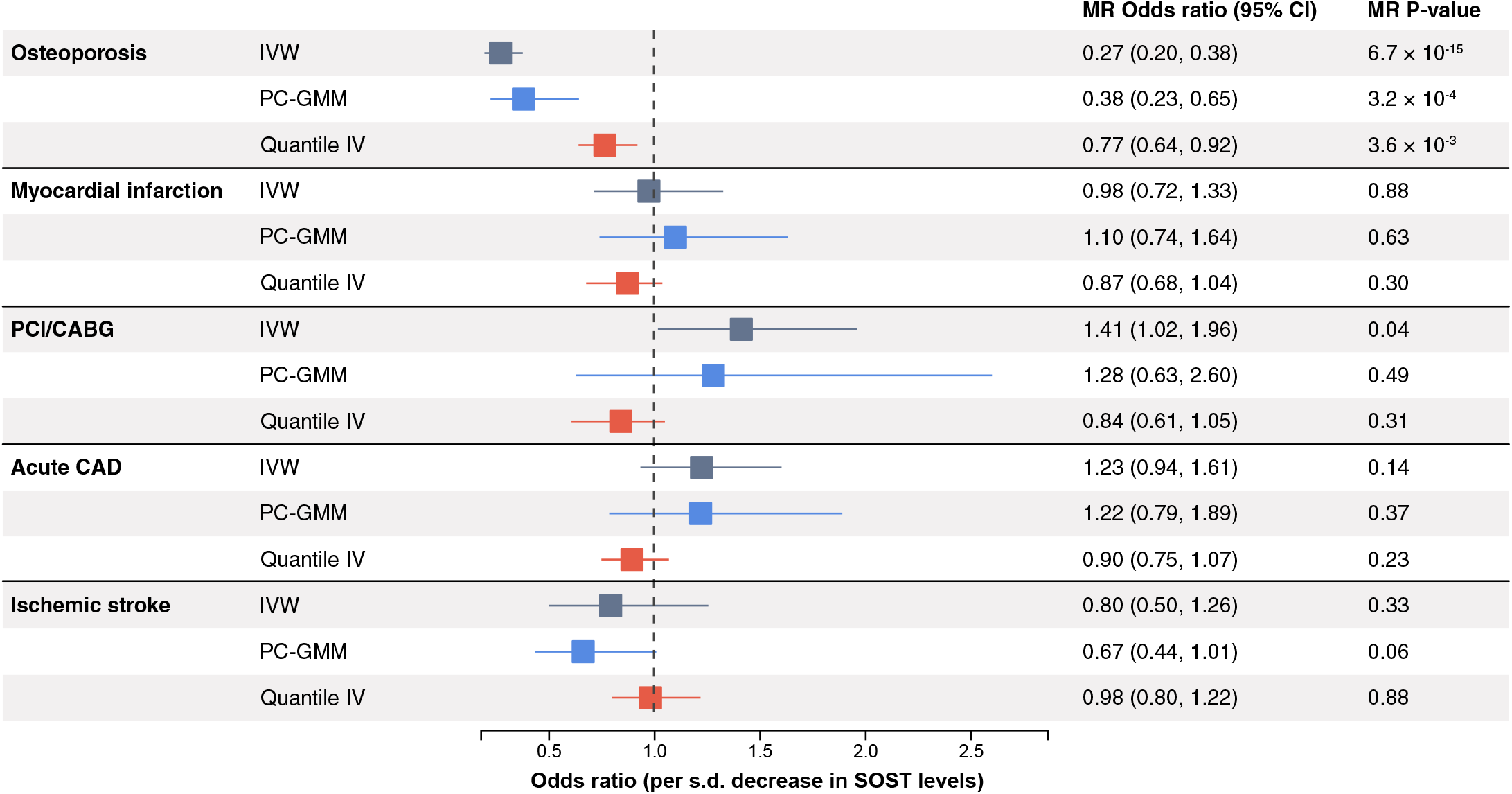
Mendelian randomization estimates of a 1 s.d. decrease in circulating sclerostin levels on osteoporosis and cardiovascular diseases accounting for direct effects by rs113533733 in the UK Biobank.

## 4 Discussion

This study introduces Quantile IV, a novel nonparametric MR estimator that offers computational stability while introducing few statistical assumptions. This innovation is particularly relevant given the limited exploration of nonparametric IV estimators in MR contexts. We evaluated Quantile IV across many realistic MR scenarios and applied it to study the causal effect of circulating sclerostin inhibition on heel bone mineral density, osteoporosis and cardiovascular diseases. Compared to DeepIV, another nonparametric IV estimator, Quantile IV consistently showed lower error and greater stability in all simulations. To our knowledge, nonparametric IV estimators have scarcely been tested and applied in MR. He *et al*. previously proposed DeLIVR, which we consider to be a semi-parametric relaxation of the fully nonparametric estimator [14]. We have confirmed that DeLIVR performs better and more consistently than DeepIV in our simulations. However, we note that this superiority comes at the cost of restrictions on the functional form of the causal effect and on imposing the additional assumption that the linear model with homoscedastic errors holds for the first stage. These two conditions were fulfilled in our simulation study favoring DeLIVR over DeepIV and Quantile IV which do not make these simplifying assumptions. However, Quantile IV consistently demonstrated robust performance with minimal error across all the tested scenarios. These scenarios probed various elements such as the impact of differing sample sizes, the strength of the IVs, the shape of the causal effect, the degree of confounding influences, and the total number of IVs employed. Quantile IV’s performance varied more in simulations with the smallest sample size (*n* = 10, 000). The only scenario where DeLIVR significantly outperformed Quantile IV is when the number of IVs was set to the largest number (100 independent IVs), but we reiterate that the simulation favored DeLIVR as the linearity and constant error assumptions held. A limitation of Quantile IV, as a machine learning-based estimator, is its lack of direct methods for uncertainty quantification. We used a bootstrap aggregation (bagging) strategy to address this limitation allowing us to construct confidence intervals and compute P-values for the hypothesis that the ATE (or CATE) is null. The bagging confidence intervals had good coverage of the true value on average, but we did notice some localized miscoverage despite low estimation error. We attribute this finding to the neural network regularization and weight initialization which may favor null effects resulting in conservative estimates when the ATE is close to zero. The false positive rate was well controlled in our simulations, never exceeding the nominal level.

The causal effect of inhibiting sclerostin on bone and cardiovascular health has been predicted using MR in the past [23, 27, 31]. However, the results from previous MR studies are conflicting and we opted to revisit the question using updated MR estimators, support from finemapping and colocalization analyses and circulating sclerostin measurements from the UK Biobank Pharma Proteomics Projects. To briefly summarize previous work, Bovijn *et al*. used IVs at the *SOST* locus that were ascertained based on their effect on bone mineral density. Using MR, they recapitulated the protective effect of sclerostin inhibition on osteoporosis and fracture risk, and estimated an increase of 18% in the odds of MI per 0.09 g/cm^2^ of bone mineral density (*P* = 0.003) [23]. The second MR study by Holdsworth *et al*. selected genetic variants at the *SOST* locus that were eQTLs of *SOST* in arterial or heart tissue and associated with bone mineral density [27]. These variants were reported to not be associated with cardiovascular outcomes including MI (*P* = 0.73) and CAD (*P* = 0.73) in CARDIoGRAMplusC4D. Another study by Zheng *et al*. conducted a GWAS meta-analysis of circulating sclerostin including 33,961 individuals from 9 cohorts. A total of 18 conditionally independent variants were associated with circulating sclerostin and the authors have conducted MR based on all of the variants (including *trans* effects) and a subset of *cis* acting variants. The *cis*-MR analysis from Zheng *et al*. suggests an increased risk of MI with an OR of 1.35 (*P* = 0.04) per 1 s.d. lowering of sclerostin levels. Faced with these contradictory results, we first investigated the possibility of bias due to LD with variants that could influence cardiovascular disease risk independently from sclerostin. Using finemapping, we were able to infer two credible sets explaining the *cis*-regulatory signal of circulating sclerostin levels. The variants in the circulating sclerostin credible sets colocalized with the osteoporosis and heel bone mineral density credible sets, but not with MI or other cardiovascular diseases. This result suggests that previous MR estimates may have been biased due to the presence of LD with cardiovascular risk variants that act independently from the modulation of circulating sclerostin levels.

We then conducted *cis*-MR analyses to estimate the effect of a 1 s.d. reduction in circulating sclerostin on heel bone mineral density, osteoporosis and cardiovascular outcomes. In accordance with the well-known clinical effect of pharmacological sclerostin inhibition [21, 22], our MR estimates showed that a genetically predicted reduction in circulating sclerostin levels leads to an increase in heel bone mineral density and a decrease in the risk of osteoporosis. There was evidence of sex differences with a larger increase in bone mineral density for a same reduction in sclerostin levels in women compared to men (0.15 vs 0.09). This is concordant with the difference observed comparing the ARCH trial of postmenopausal women to the BRIDGE trial of men where monthly injection of 210mg of romosozumab increased lumbar spine bone mineral density by 13.7% vs 12.1% and total hip bone mineral density by 6.2% vs 2.5% [21, 22]. In our study, the magnitude of the Quantile IV estimate for the effect of sclerostin inhibition on heel bone mineral density and osteoporosis were 4 to 5 times smaller than the parametric estimates from IVW and PC-GMM. This effect could be due to the linear extrapolation of small genetic effects (*e*.*g*. allelic effects of < 0.1s.d.) to predict the effect of a comparatively larger 1 s.d. decrease in circulating sclerostin. MR estimates are often larger than effects estimated in randomized controlled trials and the difference is typically attributed to the comparison of lifelong exposure effects compared to the effect of short-term interventions after disease onset [32]. Whether violations of parametric assumptions contribute to the inflation of effect estimates in real world settings is unclear. To better contextualize these findings, it would be interesting to compare the MR estimate to the pharmacological effect of romosozumab. However, to maximize statistical power in our study, we estimated causal effects on estimated heel bone mineral density which is different from clinical measurements used in clinical trials which rely on total hip and lumbar spine bone mineral density acquired via dual-energy X-ray absorptiometry. The effect of anti-sclerostin monoclonal antibodies on circulating sclerostin levels has not been assessed in clinical trials further hampering direct comparisons with MR. Finally, our MR estimates accounting for possible bias due to LD were concordant with colocalization analyses and results from large CAD genetics consortia and found a null relationship between genetically predicted sclerostin levels and ischemic cardiovascular diseases.

In this study, we proposed a new MR estimator, Quantile IV, and demonstrated its performance in simulation models. Our estimator makes few modeling assumptions when compared to traditional methods and it allows for non-linearity and effect heterogeneity. Unlike other MR estimators, Quantile IV allows the estimation of conditional average treatment effect without specifying an interaction model, which is an important tool to assess treatment response heterogeneity. This could be used, for example, to better predict whether subgroups of patients may benefit more from a medical intervention. Our work only covered a small subset of the literature on machine learning IV estimation. We focused on estimators from the nonparametric IV estimation literature stemming from the work of Newey and Powell [18]. There are alternative approaches that have yet to be evaluated in MR including methods based on kernel instrumental variable regression, deep generalized method of moments or linear IV models with learned representations [33–35]. How these methods compare to Quantile IV and other MR estimators remains unknown. Despite good overall performance, Quantile IV is a computationally intensive method, especially if confidence intervals need to be estimated via bootstrapping. This could be alleviated by using more efficient forms or bootstrapping or deep probabilistic models which could be considered in future work. Another limitation of any nonparametric estimator is that it requires access to individual level data.

We applied our estimator to tackle a challenging medical question, namely, to predict the effect of sclerostin inhibition on bone and cardiovascular health. We opted for a two-sample design within the UK Biobank to reduce the risk of weak instrument bias due to statistical sampling. Our MR study and colocalization analyses suggest that inhibiting sclerostin increases heel bone mineral density and reduces the risk of osteoporosis in the general population. This result is concordant with the findings of clinical trials of romosozumab, a pharmacological sclerostin inhibitor. Our MR and colocalization analyses suggest that inhibition of sclerostin will not lead to an increased risk of ischemic cardiovascular diseases. Because of the limited number of observed cardiovascular outcomes in our study, it is possible that a small causal effect would remain undetected due to low statistical power. However, a false negative finding is unlikely because the lead sclerostin pQTLs were not significantly associated with MI or CAD in the largest available dataset of GWAS summary statistics from the CARDIoGRAMplusC4D consortium. The numerical increase in cardiovascular events observed in individuals treated with romosozumab in clinical trials could be explained by off target effects that are independent from the modulation of sclerostin and not captured by genetic studies.

Our study not only introduces a promising MR estimator but also provides new insights into the effects of sclerostin inhibition on bone and cardiovascular health. These findings contribute to the broader understanding of the on-target effects of sclerostin inhibition and the potential of MR to study drug safety and efficacy.

## 5 Methods

### 5.1 Study population

The UK Biobank is a densely phenotyped population cohort of 500,000 participants that have been genotyped and imputed [36]. At the recruitment visit, UK Biobank participants undergo a thorough assessment with health questionnaires (touchscreen and verbal interview), blood and urine biomarker panels and physical measurements including ultrasound bone densitometry. Linkage to national health system hospitalization and death records further enable the algorithmic definition of many diseases including acute cardiovascular events (Supplementary Methods, Supplementary Table A6). A subset of 46,673 randomly selected participants enrolled in the UK Biobank Pharma Protemics Project have high throughput proteomics data measuring around 3,000 circulating proteins using the Olink platform. The current study is based on a subset of the UK Biobank cohort described in more details in the Supplementary Methods.

### 5.2 Genetic association analyses

To identify genetic variants associated with circulating sclerostin levels (pQTLs), we conducted a genetic association analysis of 1,449 common (MAF *≥* 1%) biallelic genetic variants at the *SOST* locus in 42,830 UK Biobank participants with available sclerostin measurements. The circulating sclerostin measurements are taken from high throughput proteomics measurements of circulating proteins (Supplementary Methods) [6]. We defined the *SOST* locus using the gene boundaries and including 400kb padding upstream and 200kb padding downstream. The final coordinates of the locus on the GRCh37 reference build are chr17:41,631,099-42,236,156. We used Plink v2.00a2LM AVX2 Intel (25 Oct 2019) using the generalized linear model `(--glm)` option implementing linear and logistic regression for association testing. The association statistics (*i*.*e*. estimated coefficients and standard errors) were subsequently used for MR estimation using parametric models. We used the same procedure to estimate the effect of genetic variants at the *SOST* locus on the outcomes considered in the MR study. We used linear regression for heel bone mineral density and logistic regression for osteoporosis, PCI/CABG, MI, acute CAD and ischemic stroke. All the genetic association models were adjusted for age at baseline, sex and the first 5 principal components to adjust for residual population structure.

### 5.3 Gene expression and cardiovascular disease summary statistics

We used data from the CARDIoGRAMplusC4D consortium to evaluate genetic associations from a well-powered study. We used summary statistics from a GWAS meta-analysis considering genetic variants imputed using the 1000 Genomes project data and including 60,801 cases of CAD [37]. We also used data from the updated meta-analysis from CARDIoGRAMplusC4D that includes 10,801 additional CAD cases from the UK Biobank [38]. We consider this dataset for colocalization and association analyses, but not for the MR analyses because Quantile IV requires individual level data, as do most nonlinear MR estimators.

We investigated the overlap between the genetic regulation of sclerostin circulating protein level and gene expression quantitative trait loci (eQTLs) in aorta and tibial artery tissues in GTEx V8 [39]. We selected these tissues because they are the ones with the most *SOST* expression and they are plausible candidates to explain a cardiovascular impact of sclerostin. We note that bone tissue is not included in GTEx hampering our ability to identify bone eQTLs of sclerostin.

### 5.4 Finemapping and colocalization analyses

Finemapping is used to infer credible sets of genetic variants that, under the model assumptions, will include the true causal variant at a specified probability level. We used the “Sum of Single Effects” statistical model (SuSiE) which considers the sum of regression models with a single non-zero effect for finemapping. This approach can accommodate multiple causal variants within a region and allows the computation of variant posterior inclusion probabilities (PIPs) for every variant [25]. We used the implementation from the “susieR” R package (v0.12.35).

Colocalization analysis compares the estimated association between genetic variants and a pair of traits to infer the presence or absence of shared causal variants. The “coloc” model estimates the posterior probability of five mutually exclusive hypotheses. H_0_ is the hypothesis that there are no causal variants, H_1_ that there is only a causal variant for trait 1, H_2_ that there is only a causal variant for trait 2, H_3_ that there are causal variants for both traits and that they are distinct and H_4_ that there is a shared causal variant for both traits. The original publication only accounted for a single causal genetic variant per association signal [40], but this assumption was subsequently relaxed [26]. To account for multiple causal variants, finemapping is first used to derive credible sets, and pairwise colocalization between credible sets for the two traits is tested. We use this approach in our study in instances where we were able to infer credible sets with coverage ≥ 85%. When unable to infer credible sets, we assumed a maximum of a single causal variant per trait. We used the “coloc” R package (v5.2.2) to conduct all of the colocalization analyses (“coloc.abf” and “coloc.susie” functions) with the default prior values and LD matrices computed in the subset of UK Biobank participants that passed our genetic quality control. We followed the recommendations from the “susieR” package authors and verified that the, λ statistic had low values and that the kriging plot did not have outlier variants to ensure a adequate matching between the LD matrix and summary statistics and to detect allele flips.

### 5.5 Causal assumptions

The MR estimators used in this study rely on the three main IV assumptions (Supplementary Figure A17) [41]. We denote the exposure of interest as *X*, the outcome as *Y*, the observed covariables as *W* and the instrumental variables as *Z*. The first assumption (IV1), *relevance*, states that the instrumental variable is not independent from the exposure 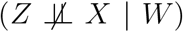. The second assumption (IV2), assumes *unconfoundedness* of the IV meaning that the IV is independent from unobservable confounders of the exposure–outcome relationship (*Z* ╨ *U* | *W*). Third, we assume the *exclusion restriction* (IV3) also commonly known as the “no horizontal pleiotropy” assumption in the MR literature. This assumption requires that the effect the IV exerts on the outcome is exclusively through the modulation of the exposure (*Z* ╨ *Y* | *X, W*). Common violations of this assumption include direct pleiotropic effects of the IV on the outcome and, more perniciously, effects due to LD with a variant influences the outcome independently from the exposure. Many attempts to relax these assumptions rely on the identification of subset of IVs with homogeneous effects, the inference of statistical structure in the causal effects or on the estimation of the mode of the causal effect (*e*.*g*. [10, 42, 43]). These approaches are useful, but they are not suitable for use in *cis*-MR because a single set of correlated candidate IVs is used hampering the estimation of modes, the inference of latent statistical structure or the detection of outliers.

In our MR study investigating the effect of circulating sclerostin levels on bone and cardiovascular health, we were able to confirm the relevance assumption by observing a strong association between our IVs, rs6416905 and rs66838809, and circulating sclerostin levels (*P* = 1.49 *×* 10^−18^ and *P* = 1.82 *×* 10^−16^ respectively). The *F* statistic for these two IVs was 60. The most plausible violation of the unconfoundedness assumption is population structure and we mitigated this risk by using a genetically homogeneous subset of participants within the UK Biobank and further adjusting all the MR estimates for the first 5 genetic principal components. Finally, since we only considered genetic variants associated with circulating sclerostin levels at the *SOST* locus, it is plausible that the observed effects are due to the modulation of sclerostin levels and not via other pathways. The risk of violations of the exclusion restriction assumption mostly arises from bias due to LD with other variants that may influence the considered outcomes. We use colocalization as an analytical approach to confirm that the causal variants underpinning genetic associations with the exposure and outcome are shared. We also conducted sensitivity analyses adjusting for genetic variants that are likely to have direct effects on the outcome when there was evidence for direct effects.

### 5.6 Two-sample parametric MR

We estimated the causal effect of circulating sclerostin levels on heel bone mineral density, osteoporosis, MI, acute CAD, PCI/CABG and ischemic stroke using genetic variants at the *SOST* locus as IVs and using parametric MR models suitable for the *cis*-MR context. Specifically, we used the IVW and PC-GMM estimators. The IVW is a weighted average of the ratio estimates of the IVs where the weights are proportional to the precision of the ratio estimates [44]. The PC-GMM estimator was designed for the *cis*-MR setting and uses a principal component analysis of a weighted LD matrix accounting for instrument strength and the precision of the effect estimates on the outcome [45]. The principal components from this decomposition are then used as IVs using generalized method of moments for estimation [29]. We used the robust standard errors accounting for overdispersion in the current study. For the IVW and PC-GMM estimators, we used the implementation from the R “MendelianRandomization” package (v0.9.0).

### 5.7 Nonparametric IV estimation

We first consider the model introduced by Hartford *et al*. when describing the DeepIV estimator [20]:

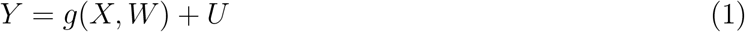

In our context, *Y* represents the outcome, *X* denotes the exposure, *W* encompasses observed confounders. The latent variables *U* account for the unobservable factors that may affect *Y, X* and *W*. It enters the model additively. Here *g*(*·*) is some unknown and potentially nonlinear function of both *X* and *W*. We further introduce an instrumental variable (*Z*) that satisfies the IV assumptions (IV1-3, Supplementary Figure A17).

The goal is to estimate the conditional average treatment effect (CATE) *η*(*w*) defined as

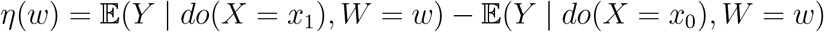

Under the do operator and the assumption of the model (1), we have

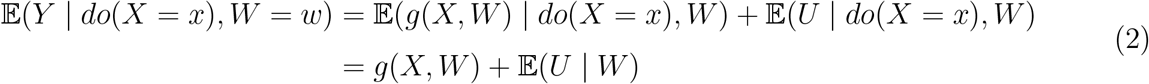

We can estimate the CATE by estimating the *h*(*·*) function defined as

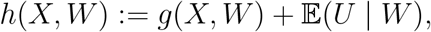

since the conditional expectation of the confounder given the covariates will not influence the estimation of contrasts such as the CATE: *η* (*w*) = *g*(*x*_1_, *w*) − *g*(*x*_0_, *w*) = *h*(*x*_1_, *w*) − *h*(*x*_0_, *w*).

Note that if we additionally assume that 𝔼 (*U* | *W*) = 0, then *h*(*X, W*) directly characterizes the conditional effect of an intervention of *X* on *Y*.

To estimate *h*(*·*), DeepIV uses the following result

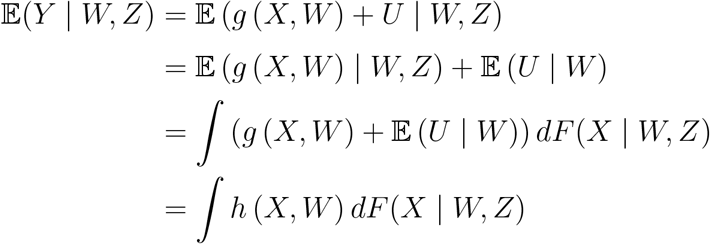

Based on this result, Hartford *et al*. suggest estimating *h*(*·*) by solving the following optimization problem:

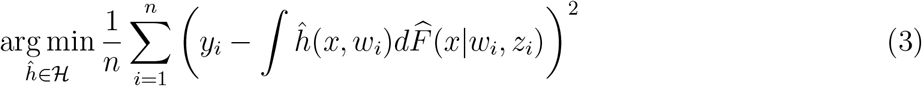

Which is found using a two-stage procedure. The first stage uses a treatment network to estimate the conditional cumulative distribution function 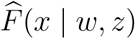 using any statistical or machine learning model such as the mixture density network which was initially suggested; The second stage then samples conditional exposure values from the first stage network and relates these samples to the observed outcome to estimate *h*(*x, w*).

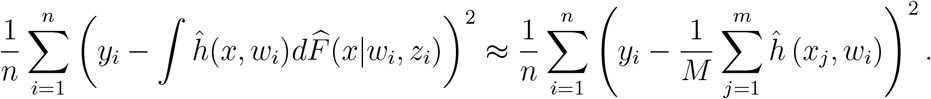

where *x*_*j*_ are samples from the estimated cumulative density function 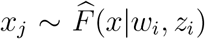. This step is akin to any supervised learning task and can be done using a feedforward neural network. We note that this procedure is analogous to the two-stage least squares procedure that is a conventional estimator in instrumental variable analysis.

This procedure relaxes the parametric assumptions of conventional MR methods and only assumes that the confounder enters additively in the model. However, the sampling step needed to train the second stage introduces additional stochasticity in the training process and may limit performance in MR [14].

### 5.8 Quantile IV algorithm

The current method, Quantile IV, proposes replacing the density estimation in the first stage of the DeepIV procedure by a quantile regression which eliminates the need for sampling and simplifies the optimization. Quantile IV is a specific instantiation of DeepIV and is an equally valid estimator while performing advantageously in realistic settings.

We now describe the estimation strategy for Quantile IV. In the first stage, we estimate *K* evenly spaced conditional quantiles of the exposure given the instruments using a neural network trained using the quantile loss. Specifically, we wish to estimate a fixed number of evenly spaced conditional *τ*-th quantile of *X* given *W* and *Z* with 0 < *τ* < 1.

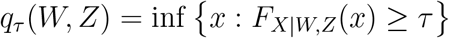

where *F*_*X*|*W,Z*_(*x*) = *P* (*X* ≤ *x*|*W, Z*) is the conditional distribution function of *X*. We know that the expectation of any function *f*(*X*) of some random variable *X* can be related to the quantile using the following relation: 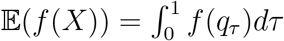, where q_*τ*_ represents the *τ*-th quantile of *X*. In our context, this provides us a method to approximate the integral ∫ *h* (*X, W*) *dF* (*X* | *W, Z*) using *K* conditional quantiles 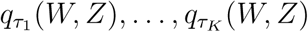. Partition (0, 1) using *K* +1 even-spaced points 0 = *a*_0_ < *a*_1_ < *·· ·* < *a*_*K*_ = 1 where *a*_*k*_ = *k*/*K* and choose quantiles 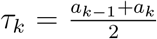 for *k* = 1,…, *K*. We have

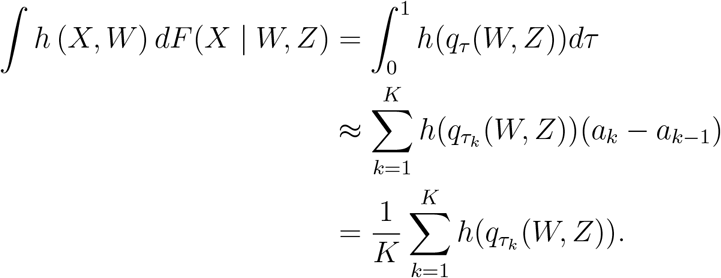

To learn quantiles q*τ*_*k*_ (*w, z*) for *k* = 1,…, *K*, we use a neural network parametrized by a set of weights and biases denoted as *ϕ* and with a *K*-dimensional output layer ***f*** (*w, z* ; *ϕ*) : *W × Z* → *X*^*K*^. Let ***f*** = (***f*** ^(1)^,…, ***f*** ^(*K*)^), where ***f*** ^(*k*)^ is the k-th element of ***f***. The quantile loss estimates conditional quantiles [46] and can be expressed as:

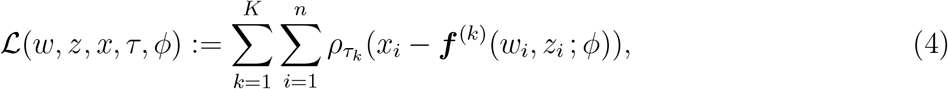

where *ρ τ* (*u*) = (*τ* − 𝕀 [*u ≤* 0])*u*. Hence *K* conditional quantiles 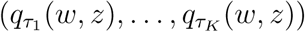 can be simultaneously estimated by ***f*** (*w, z* ; *ϕ*). In the first stage of Quantile IV, we train a neural network to minimize this quantile loss solving:

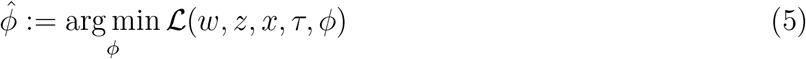

The key insight of the method is that these *K* quantiles divide the conditional distribution of the exposure into equally probable regions and allow us to replace the sampling step in DeepIV by a simple average over these conditional quantiles as the input to the second stage regression. Under this first stage model and using a second neural network *h* : *X × W* ⟶ *Y* to estimate the IV regression function, Equation (3) becomes

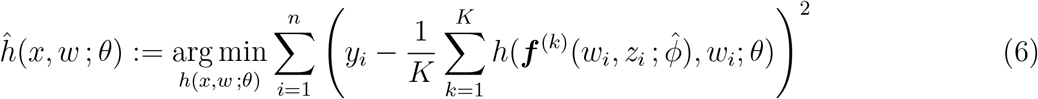

The optimization of the weights and biases (denoted by *θ*) of this neural network can be achieved using conventional gradient-based optimizers (*e*.*g*. Adam [47]).

#### 5.8.1 Neural network quantile regression

In our formulation of Quantile IV, we use a standard feedforward neural network trained with the quantile loss to estimate the conditional quantiles of the exposure given the IVs. This approach may pose problem because it does not strictly constrain the learned conditional quantiles to be monotonically increasing, even though the quantile loss encourages the estimation of quantiles satisfying this property. For example, there is no constraint in the model forcing the 90% quantile to be smaller than the 95% quantile which is required by the definition of quantiles. This problem is called the “quantile crossing” problem and Moon *et al*. have proposed a neural network model and training procedure called the Noncrossing Multiple Quantile Regression with neural networks (NMQN) to address it [48]. We have implemented the NMQN and tested the impact on Quantile IV estimates. We observed that the estimated conditional quantiles are slightly more spread out when comparing the NMQN to the naive implementation, but there was an increase in the average mean squared error in preliminary analysis of the simulation study prompting us to rely on the naive implementation. The option to use NMQN remains available in the implementation.

### 5.9 Quantile IV implementation and estimation

We implemented the Quantile IV estimator using the Python programming language and the PyTorch framework (v1.13.0, https://pytorch.org/). Our implementation is publicly available online as part of our *ml-mr* Python package (https://github.com/legaultmarc/ml-mr). The Quantile IV estimator is effectively composed of two neural networks and our implementation allows setting the corresponding hyperparameters including the number of layers, number of hidden units and learning rate. To avoid overfitting, we use a sample splitting strategy which randomly selects 20% (by default) of the samples to be used as a validation dataset. Training is based strictly on the training dataset, but the validation dataset is used to stop training when there are no further improvements on the validation loss, a strategy known as early stopping. Models can be trained using either the CPU or specialized hardware such as Graphical Processing Units (GPUs). In practice, because the neural networks used in Quantile IV are relatively small, we have not observed significant performance improvements when training on GPUs. Reasonable default values (Supplementary Table A7) for all of the hyperparameters were selected based on our experimentation on both real and simulated data during model development. We used these default values from our software implementation unless otherwise specified.

### 5.10 MR Simulation study

We conducted a simulation study to assess the performance of nonparametric IV estimators in the context of MR. To achieve this, we selected simulation parameters covering a range of plausible MR settings. We vary the structural relationship between the exposure and outcome between a linear, J-shaped (quadratic) and threshold effect. These forms of nonlinearity are the most relevant to medical applications and are commonly studied in epidemiology. We use the specific parametrization from previous simulations of nonlinear MR as summarized in Table 1 [12]. We vary the sample size between 10,000 individuals to 100,000 corresponding to plausible modern sample sizes for small and large genetic studies. We use the heritability of the exposure explained by the instrumental variables 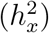 to vary the strength of the genetic IVs. We simulate traits with 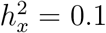 corresponding to traits with a small fraction of the variance explained by genetic factors, traits with moderate heritability 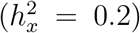 and traits with high heritability 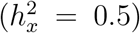. For reference and context, using the phenome-wide heritability browser based on the UK Biobank data (https://nealelab.github.io/UKBB_ldsc/h2_browser.html), we observed that body mass index had an estimated *h*^2^ = 0.25, systolic blood pressure had *h*^2^ = 0.15, heel bone mineral density (right) had *h*^2^ = 0.33 and forced expiratory volume in 1-second had *h*^2^ = 0.43. We emphasize that in our study, we only consider the heritability explained by the instrumental variable which is lower than the genome-wide SNP heritability.

To simulate genetic variants to be used as IVs, we follow the procedure described in Sulc *et al*. [11]. Briefly, we sample allele frequencies *p*_*i*_ ∼ Beta(1, 3) and draw the number of alternative alleles following a binomial distribution with two draws. We then standardized the genotypes and assigned the effects using the baseline LDAK heritability model as (*β*_*i*_ ∼ 𝒩 (0, *p*_*i*_(1 − *p*_*i*_)^*-*0.25^) and rescaling to reach the desired heritability. The exposure and outcomes are then simulated as

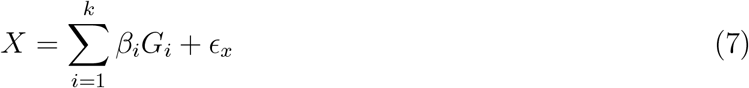

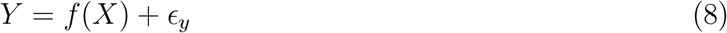

where *G*_*i*_ are the simulated standardized genotypes, *f*(*X*) is the simulated structural relationship between the exposure and the outcome and with

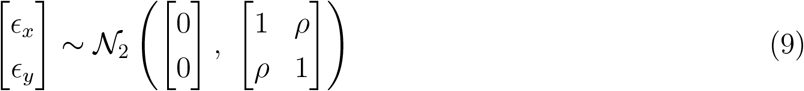

The formulation for the errors implicitly models the effect of unmeasured confounding and follows the simulation model from He *et al*. [14]. The advantage of this formulation is that it provides a convenient way of varying the strength of the latent confounding variable using a single simulation parameter (*ρ*). When varying the simulation parameters, we hold the others fixed at a reference value indicated in Table 1 (bold values).

### 5.11 Estimating treatment effects and their confidence intervals

The Quantile IV algorithm estimates the IV regression function *ĥ*(*x, w*; *θ*). From this fitted model, an estimator for the CATE is given by:

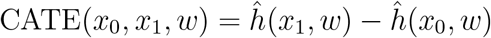

where we dropped parameters for notational convenience. Similarly, an estimate of the ATE is obtained by averaging the CATE over the empirical data distribution.

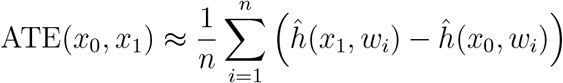

To obtain confidence intervals, we rely on bootstrapping [49]. We resample the dataset with replacement and refit Quantile IV for every one of the *B* bootstrap resamples. This yields a bag of IV regression functions 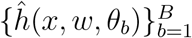. Bootstrapping in this way first allows us to ensemble the predictions to obtain a bagging estimator for the CATE (and consequentially the ATE):

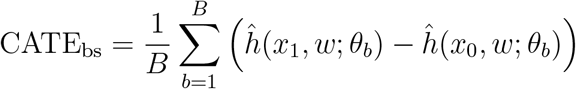

A confidence interval at the 1 − *α* coverage level is obtained by selecting the *α*/2 and 1 − *α*/2 quantiles of the bootstrap estimates of the CATE. We derive P-values from these confidence intervals by assuming asymptotic normality to avoid the high computational cost of computing bootstrap P-values. Interaction P-values are computed using one way ANOVA of the bootstrap estimates of the CATEs at varying levels of the conditioning variable.

### 5.12 Evaluation of the Quantile IV estimator

In simulation scenarios, we had access to the true causal relationship between the exposure and outcome *g*(*X, W*). In the simulation, the conditional expectation of the confounder given the covariates is 0 meaning that the estimated IV function is an estimate of the interventional effect under our model (*i*.*e. 𝔼* (*U* |*W*) = 0 in Equation (2) and 𝔼 (*Y* |do(*X*),*W*) = *g*(*X, W*) = *h*(*X, W*)). Hence, we were able to assess model performance as the root mean squared error between the estimated function and the true function over a grid spanning the range of the exposure.

## Data Availability

The data used in this study is from the UK Biobank accessed under application #20168. The access procedure are described on the UK Biobank website at: https://www.ukbiobank.ac.uk/enable-your-research/apply-for-access

## 8 Acknowledgements

MAL is supported by a fellowship from the Canadian Institute of Health Research (CIHR). BJA holds a Senior Scholar Award from the Fonds de recherche du Québec: Santé.

## 9 Author contributions

MAL, BJA and JP contributed to the study design and application of the method to UK Biobank data. MAL, JH and AYY contributed to the methodological development of the Quantile IV algorithm and design the MR simulation study. All of the authors contributed to writing the manuscript and approved the final submitted version.

