## Supplementary Materials for "A novel and efficient machine learning Mendelian randomization estimator applied to predict the safety and efficacy of sclerostin inhibition"

### A Supplementary Note

#### A.1 Forward stepwise regression analysis of sclerostin pQTLs

In addition to the finemapping analysis, we also conducted a forward stepwise regression analysis where we iteratively condition on the top variant and repeat the association study. The initial association scan had identified rs9303537 as the lead variant ("T" allele $\hat{\beta} = 0.059$ 95% CI (0.045, 0.071), $P = 1.5 \times 10^{-18}$). This variant is almost perfectly correlated with the lead variant from the first credible set ($r^2 = 0.99$ in 1,000 genomes European sample). Conditioning on this top variant in the forward stepwise analysis, the variant rs66838809 was identified as the second stage top variant. The conditional association coefficient for the rs66838809 "A" allele is $\hat{\beta} = -0.078$, 95% CI (-0.091, -0.064) $P = 1.2 \times 10^{-10}$. After conditioning on these two variants, no residual signal crossed the genome-wide significance threshold. The top variant (rs75508812) had a conditional association $P = 3.3 \times 10^{-5}$. This variant passes the conservative Bonferroni correction at $\alpha = 5\%$ considering the 1,449 variants we included and can be considered significant.

### B Supplementary Methods

#### B.1 Genetic quality control

We excluded variants or individuals with a missing rate > 2%. The genetic and self-reported sex variables were compared to ensure concordance between genetic and self-reported sex and individuals with discrepancies or sex chromosome aneuploidies were excluded from the analysis dataset. To mitigate bias due to population stratification, we restricted our analysis to individuals of European ancestry because they represent the largest genetically homogeneous subgroup in the UK Biobank and excluded participants that fell outside of a manually defined region on the principal component analysis plot. Related individuals were excluded based on a kinship coefficient cutoff of 0.0884, corresponding to a relationship no closer than the 3rd degree. A total of 413,138 individuals remained after this quality control process.

#### B.2 Mendelian randomization exposure and outcome definitions

The exposure for the MR study is circulating sclerostin levels as measured using the Olink Explore high throughput proteomics platform. This platform uses the proximity extension assay technology where antibodies bind the target protein and allow hybridization of complementary affixed olignucleotides which are amplified and used to quantify protein levels. Sclerostin is assayed on this platform (protein #2527) and we extracted measurements in the normalized protein expression (NPX) format at the baseline instance. The NPX values are provided by the UK Biobank following the Olink QC and quantification process and have an approximately logarithmic interpretation (see UK Biobank resources #4654 to #4658 for additional information on the quality control and quantification) [6]. We visually confirmed that the extracted sclerostin levels had an approximately normal distribution with no outliers. A total of 42,830 individuals had available sclerostin measurements at the baseline visit and were included in our dataset accounting for our genetic quality control (Table A2).

For the MR study, we considered heel bone mineral density, osteoporosis, MI, PCI/CABG, acute CAD and ischemic stroke as outcomes. We used data from ultrasound bone densitometry which provides an estimate of heel bone mineral density in $g/cm^2$. We extracted measurements from the initial assessment visit and used the average measurement when multiple values were present for a same individual. We standardized the measurements to have a mean of 0 and standard deviation (s.d.) of 1. The reported effects in the s.d. scale can be converted back to $g/cm^2$ by multiplying by 0.14 and represent changes about the mean which is of 0.54 $g/cm^2$.

We defined the osteoporosis variable as self-reported osteoporosis (UK Biobank variable #20002) or from ICD10 codes M80 (osteoporosis with pathological fracture), M81 (osteoporosis without pathological fracture), M84.4 (pathological fracture, not elsewhere classified) or M85.9 (disorder of bone density and structure, unspecified) in the hospitalization records. We relied on hospitalization and death records to capture acute cardiovascular events using the definitions we previously developed in collaboration with physician researchers at the Montreal Heart Institute (Supplementary Table A6). Our previous definitions did not include ischemic stroke, so we relied on the definition from the UK Biobank Outcome Adjudication Group based on self-reported ischemic stroke and the codes reported in Supplementary Table A6). In addition to the coding of cases based on the diagnostic codes, we excluded individuals with self-reported MI from the controls of the MI and acute CAD variable and the individuals with self-reported ischemic stroke from the controls of the ischemic stroke variable.

### C Supplementary Tables

**Supplementary Table A1: Comparison between the mean root mean squared error between DeLIVR and Quantile IV across the different Mendelian randomization simulation scenarios and over the full range of the exposure distribution.** The P-value is from a paired t-test by simulation replicate.

| Simulation parameter | Sim. value | DeLIVR Mean RMSE (s.d.) | Quantile IV Mean RMSE (s.d.) | P-value |
|---|---|---|---|---|
| Sample size | 10,000 | 1.44 (0.30) | 1.39 (0.33) | 0.196 |
| | 50,000 | 0.98 (0.18) | 0.94 (0.23) | 0.037 |
| | 100,000 | 1.01 (0.23) | **0.86 (0.20)** | $1.1 \times 10^{-11}$ |
| Instrument strength | 0.05 | 0.88 (0.20) | 0.87 (0.22) | 0.67 |
| | 0.1 | 1.23 (0.21) | **1.13 (0.26)** | $1.7 \times 10^{-5}$ |
| | 0.5 | 0.86 (0.28) | **0.80 (0.15)** | $8.1 \times 10^{-4}$ |
| Causal relationship shape | Linear | 0.25 (0.13) | **0.20 (0.11)** | $4.4 \times 10^{-5}$ |
| | Quadratic | 0.87 (0.17) | **0.80 (0.20)** | $2.9 \times 10^{-4}$ |
| | Threshold | 0.50 (0.07) | 0.52 (0.10) | 0.076 |
| Confounding | -0.6 | 1.15 (0.22) | 1.19 (0.24) | 0.127 |
| | 0.3 | 1.06 (0.23) | **0.98 (0.23)** | $2.1 \times 10^{-4}$ |
| | 0.6 | 0.91 (0.21) | **0.75 (0.19)** | $1.3 \times 10^{-15}$ |
| Number of instruments | 2 | 1.03 (0.36) | **0.96 (0.35)** | $3.0 \times 10^{-3}$ |
| | 10 | 1.04 (0.18) | **0.96 (0.23)** | $5.1 \times 10^{-5}$ |
| | 100 | **1.01 (0.16)** | 1.30 (0.21) | $2.8 \times 10^{-35}$ |

**Supplementary Table A2: Descriptive statistics of the datasets used for the Mendelian randomization study evaluating the effect of a reduction in circulating sclerosting levels on outcomes related to bone and cardiovascular health in the UK Biobank.**

| **Exposure dataset (circulating sclerostin)** | |
|---|---|
| $n$ | 42,830 |
| Genetic Female - $n$ (%) | 22,981 (53.7%) |
| Age - Mean (s.d.) | 57.2 (8.1) |
| **Outcome dataset** | |
| $n$ | 370,218 |
| Genetic Female - $n$ (%) | 199,656 (53.9%) |
| Age at baseline - Mean (s.d.) | 56.8 (8.0) |
| Heel bone mineral density - $n$ | 211,692 |
| Mean in g/cm$^2$ (s.d.) | 0.54 (0.14) |
| Osteoporosis - $n$ cases / $n$ controls (%) | 18,937 / 351,281 (5.4%) |
| Myocardial infarction - $n$ cases / $n$ controls (%) | 19,925 / 348,722 (5.7%) |
| PCI/CABG - $n$ cases / $n$ controls (%) | 17,261 / 352,957 (4.9%) |
| Ischemic stroke - $n$ cases / $n$ controls (%) | 8 535 / 361,651 (2.4%) |
| Acute CAD - $n$ cases / $n$ controls (%) | 33,516 / 335,167 (10.0%) |

**Supplementary Table A3: Estimated genetic associations with circulating sclerostin levels for the top variants assigned to the two credible sets identified in the SuSiE finemapping analysis.**

| ID | Pos. (Chr. 17) | Alleles (ref/coded) | Sclerostin pQTL $\hat{\beta}$ (95% CI) | P-value |
|---|---|---|---|---|
| rs6416905 | 41,804,464 | A/G | -0.059 (-0.046, -0.072) | $1.49 \times 10^{-18}$ |
| rs66838809 | 41,798,621 | G/A | -0.097 (-0.074, -0.120) | $1.82 \times 10^{-16}$ |

**Supplementary Table A4: Conditional average treatment effects estimated using Quantile IV for a 1 s.d. reduction in circulating sclerostin levels on osteoporosis and heel bone mineral density.** The estimates are conditioned on different values of the covariates, specifically by conditioning on the genetic male vs female variable or varying the age at the mean (56.8 years), 1 s.d. below the mean (48.8 years) and 1 s.d. above the mean (64.8 years).

| **Outcome** | **Conditioning** | **CATE* (95% CI)** | **Interaction P-value** |
|---|---|---|---|
| Osteoporosis | Female | 0.79 (0.60, 0.95) | 0.04 |
| | Male | 0.80 (0.62, 0.96) | |
| | Age = 48.8 (mean − 1 s.d.) | 0.79 (0.61, 0.94) | 0.35 |
| | Age = 56.8 (mean) | 0.79 (0.61, 0.95) | |
| | Age = 64.8 (mean + 1 s.d.) | 0.81 (0.61, 0.98) | |
| Heel bone mineral density | Female | 0.15 (0.02, 0.26) | $2.3 \times 10^{-22}$ |
| | Male | 0.09 (-0.01, 0.18) | |
| | Age = 48.8 (mean − 1 s.d.) | 0.12 (0.03, 0.20) | 0.43 |
| | Age = 56.8 (mean) | 0.13 (0.02, 0.23) | |
| | Age = 64.8 (mean + 1 s.d.) | 0.12 (-0.02, 0.24) | |

* The CATE is reported on the odds ratio scale for osteoporosis and in standardized units (*i.e.* z-scores) for heel bone mineral density.

**Supplementary Table A5: Mendelian randomization estimates of the effect of a 1 s.d. reduction in circulating sclerostin levels on heel bone mineral density adjusting for possible direct effects by rs113533733.** The average treatment effect on heel bone mineral density for a 1 s.d. reduction in circulating sclerostin is presented.

| Estimator | ATE on heel BMD (95% CI) | P-value |
|---|---|---|
| IVW | 0.66 (0.57, 0.75) | $1.7 \times 10^{-49}$ |
| PC-GMM | 0.52 (0.18, 0.86) | $2.7 \times 10^{-3}$ |
| Quantile IV | 0.11 (0.03, 0.21) | $6.1 \times 10^{-3}$ |

**Supplementary Table A6: Algorithmic definition of the cardiovascular outcomes considered for the Mendelian randomization study.** Unless otherwise specified, codes are considered present if they are found as the primary cause of death or as the primary or secondary cause of hospitalization in the electronic records.

| Outcome | Code voc. | Code | Label |
|---|---|---|---|
| Myocardial infarction (MI) | ICD9 | 410 | Acute myocardial infarction |
| | | 412 | Old myocardial infarction |
| | | 411.0 | Postmyocardial infarction syndrome |
| | | 429.7 | Certain sequelae of myocardial infarction, not elsewhere classified |
| | ICD10 | I21 | Acute myocardial infarction |
| | | I22 | Subsequent myocardial infarction |
| | | I23 | Certain current complications following acute myocardial infarction |
| | | I25.2 | Old myocardial infarction |
| Percutaneous Coronary Intervention and Coronary Artery Bypass Graft (PCI/CABG) | OPCS | K40 | Saphenous vein graft replacement of coronary artery |
| | | K41 | Other autograft replacement of coronary artery |
| | | K42 | Allograft replacement of coronary artery |
| | | K43 | Prosthetic replacement of coronary artery |
| | | K44 | Other replacement of coronary artery |
| | | K45 | Connection of thoracic artery to coronary artery |
| | | K46 | Other bypass of coronary artery |
| | | K49 | Transluminal balloon angioplasty of coronary artery |
| | | K50 | Other therapeutic transluminal operations on coronary artery |
| | | K75 | Percutaneous transluminal balloon angioplasty and insertion of stent into coronary artery |
| Acute Coronary Artery Disease (Acute CAD) | Prev. defined outcome | MI | |
| | | PCI/CABG | |
| | ICD10 | I20.0 | Unstable angina (**for this particular instance, we only consider primary cause for hospitalization or death**) |
| Ischemic stroke | ICD9 | 434 | Occlusion of cerebral arteries |
| | | 434.0 | Cerebral thrombosis |
| | | 434.1 | Cerebral embolism |
| | | 434.9 | Cerebral artery occlusion, unspecified |
| | | 436 | Acute, but ill-defined, cerebrovascular disease |
| | ICD10 | I63 | Cerebral infarction |
| | | I64 | Stroke, not specified as haemorrhage or infarction |

**Supplementary Table A7: Default values for tunable hyperparameters of the Quantile IV estimator.** When the neural network is not specified, the default value applies for both the exposure and the outcome neural network.

| Parameter | Default value |
|---|---|
| $n$ quantiles | 5 |
| Exposure network units per layer | 128, 64 |
| Outcome network units per layer | 64, 32 |
| Activation function | Gaussian Error Linear Units (GELU) |
| Learning rate | $5 \times 10^{-4}$ |
| Minibatch size | 10,000 |
| Maximum number of epochs | 1000 |
| Weight decay ($l_2$ penalty) | $10^{-4}$ |

### D Supplementary Figures

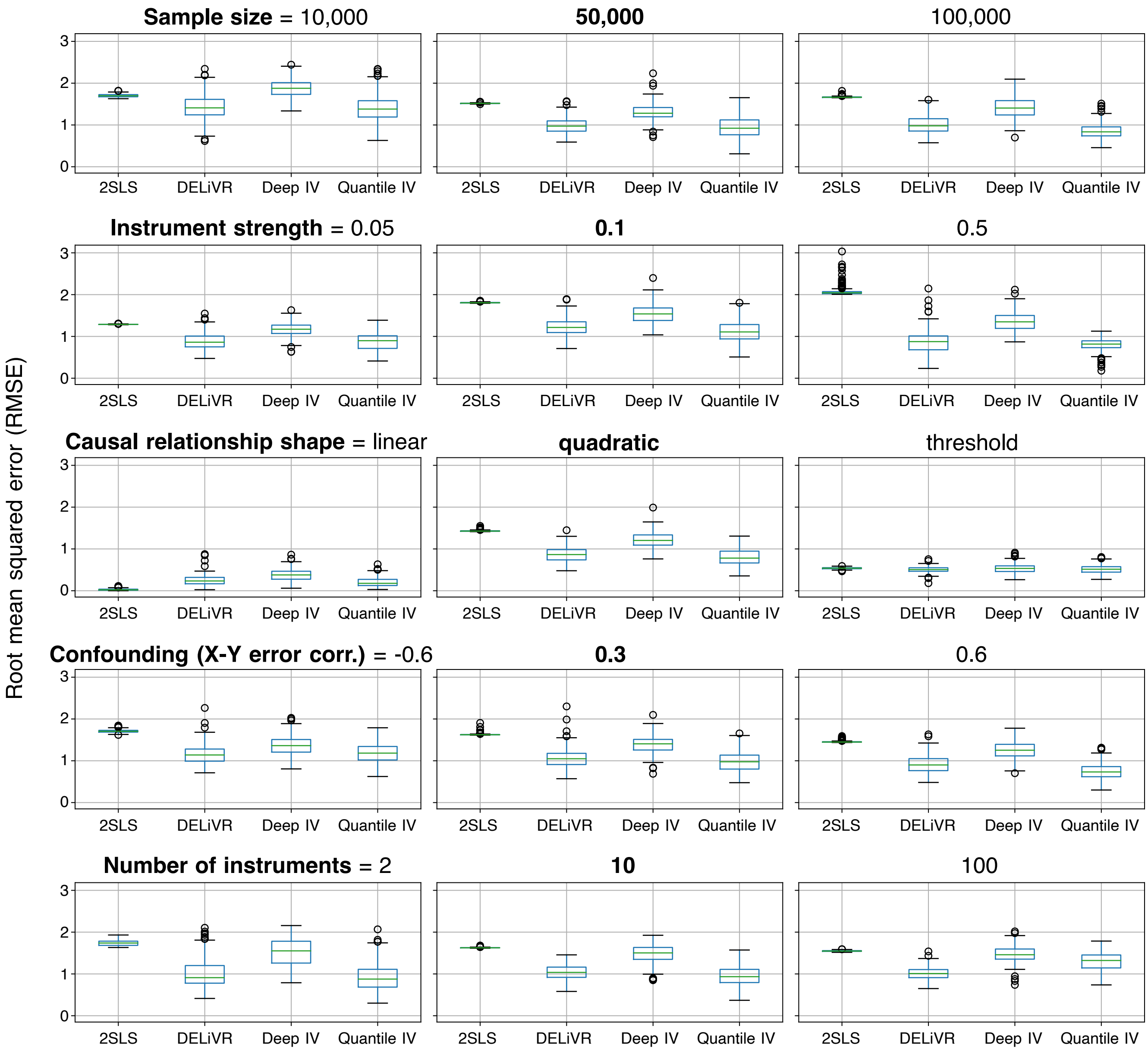


**Supplementary Figure A1: Root mean squared error between the estimated IV regression and the true causal function over a grid spanning the full range of the exposure.** The boxplot for every estimator represents variability over 200 simulation replicates. The simulation parameter values in bold correspond to the reference values. 2SLS: Two-stage least squares, DeLIVR [14], DeepIV [20], Quantile IV: proposed method.

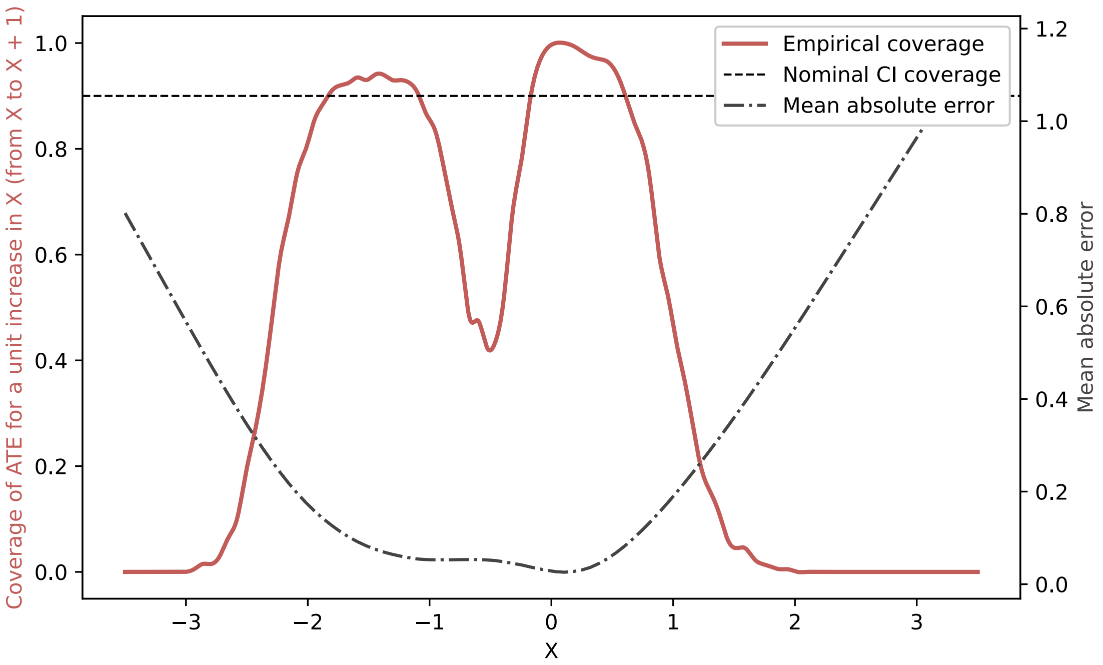


**Supplementary Figure A2: Coverage and mean absolute estimation error of the ATE for a unit increment in the exposure.** The coverage is estimated using 200 simulation replicates and 50 bootstrap iterations are used to derive the confidence intervals. The simulation scenario used for this analysis uses the baseline parameter values for every simulation parameter.

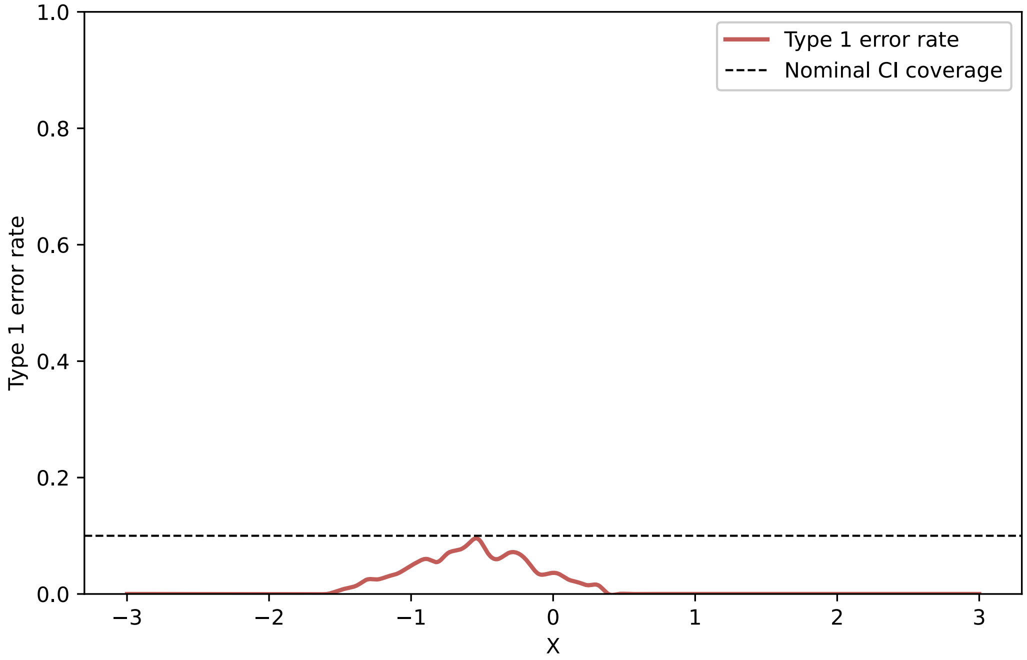


**Supplementary Figure A3: Type 1 error rate when estimating the ATE for a unit increment in the exposure across different values of the exposure.** The type 1 error rate is estimated using 200 simulation replicates and 50 bootstrap iterations are used to derive the confidence intervals. The simulation scenario used for this analysis uses the baseline parameter values for every simulation parameter except for the structural relationship where $f(X) = 0$.

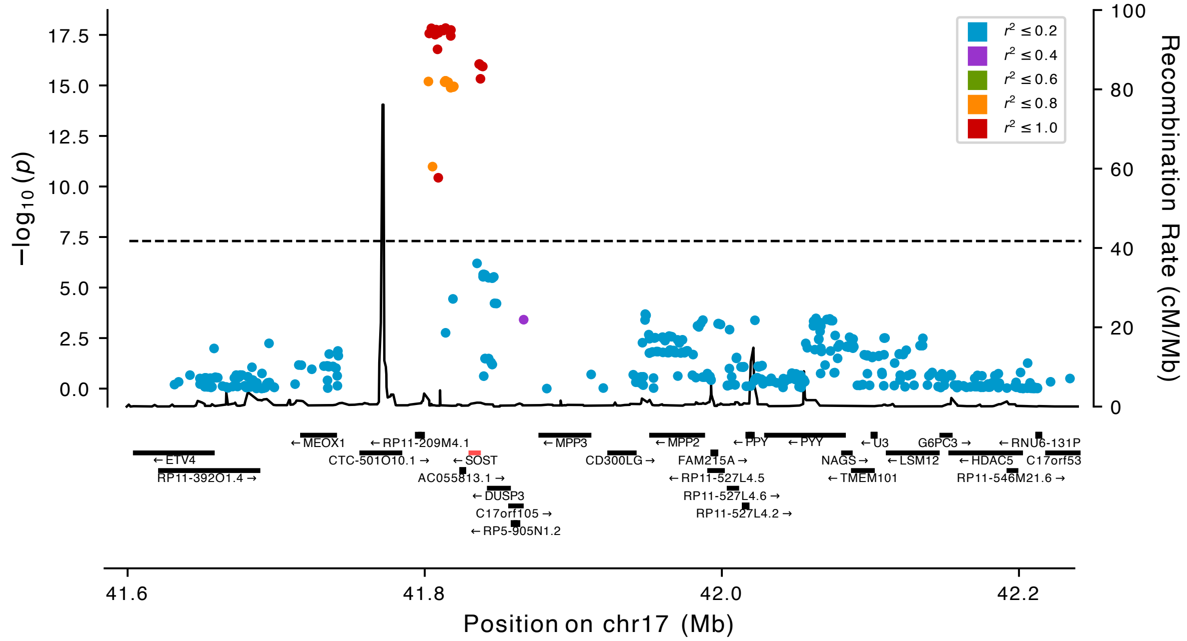


**Supplementary Figure A4: Genetic association between variants at the *SOST* locus and circulating sclerostin protein levels as measured using the Olink platform in the UK Biobank.** Genes in the region are shown below the locus plot and the *SOST* gene is highlighted in red.

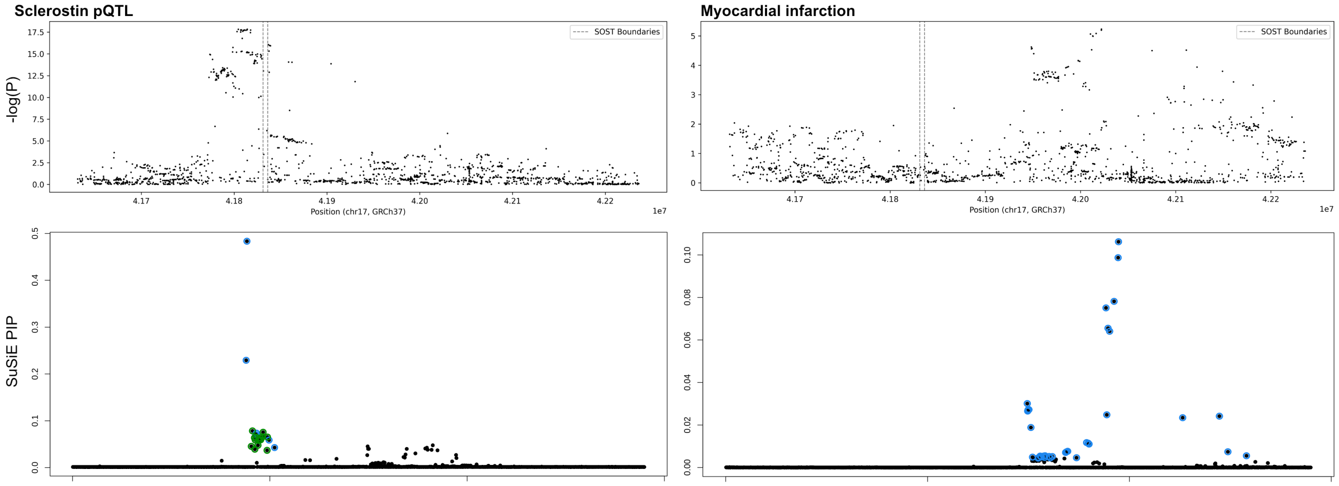


**Supplementary Figure A5: Genetic associations at the SOST locus with circulating sclerostin levels (left) and myocardial infarction (right) and variant posterior inclusion probabilities in finemapping credible sets as inferred by SuSiE.** The boundaries of the *SOST* gene are denoted by dashed lines.

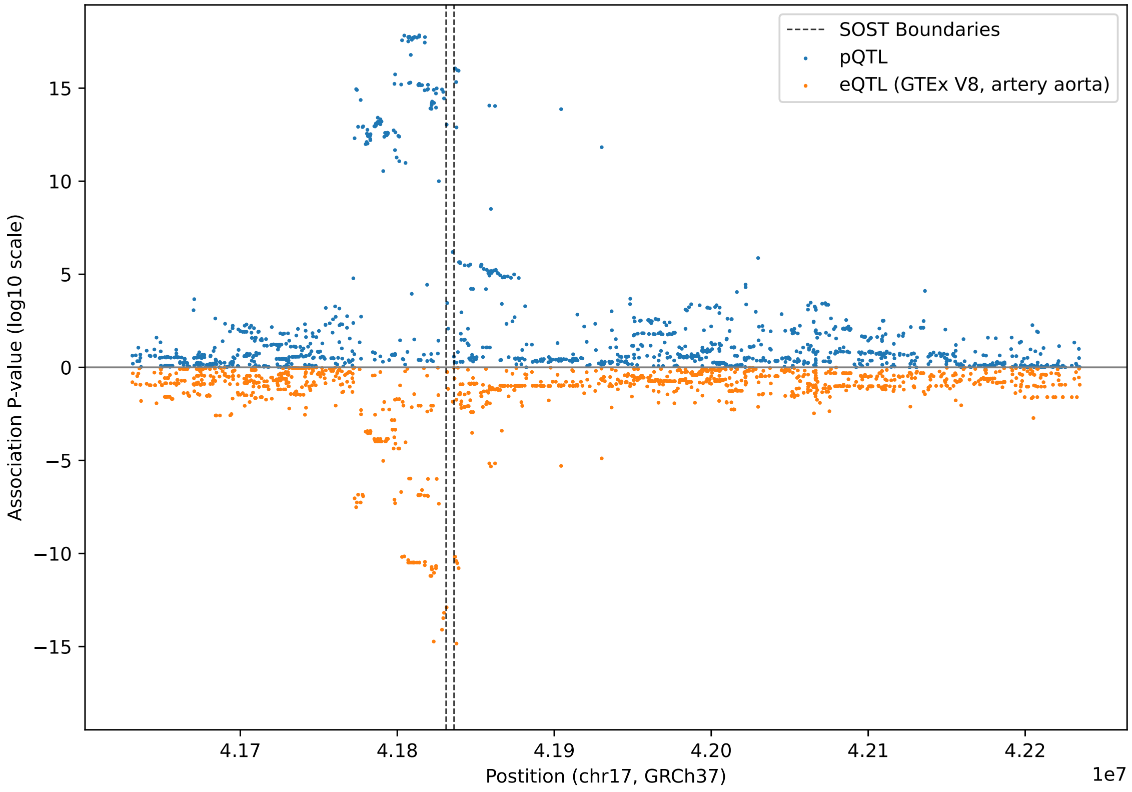


**Supplementary Figure A6: Association P-values (log scale) between genetic variants at the *SOST* locus and *SOST* expression in the aorta in GTEx V8 and circulating sclerostin levels in the UK Biobank.**

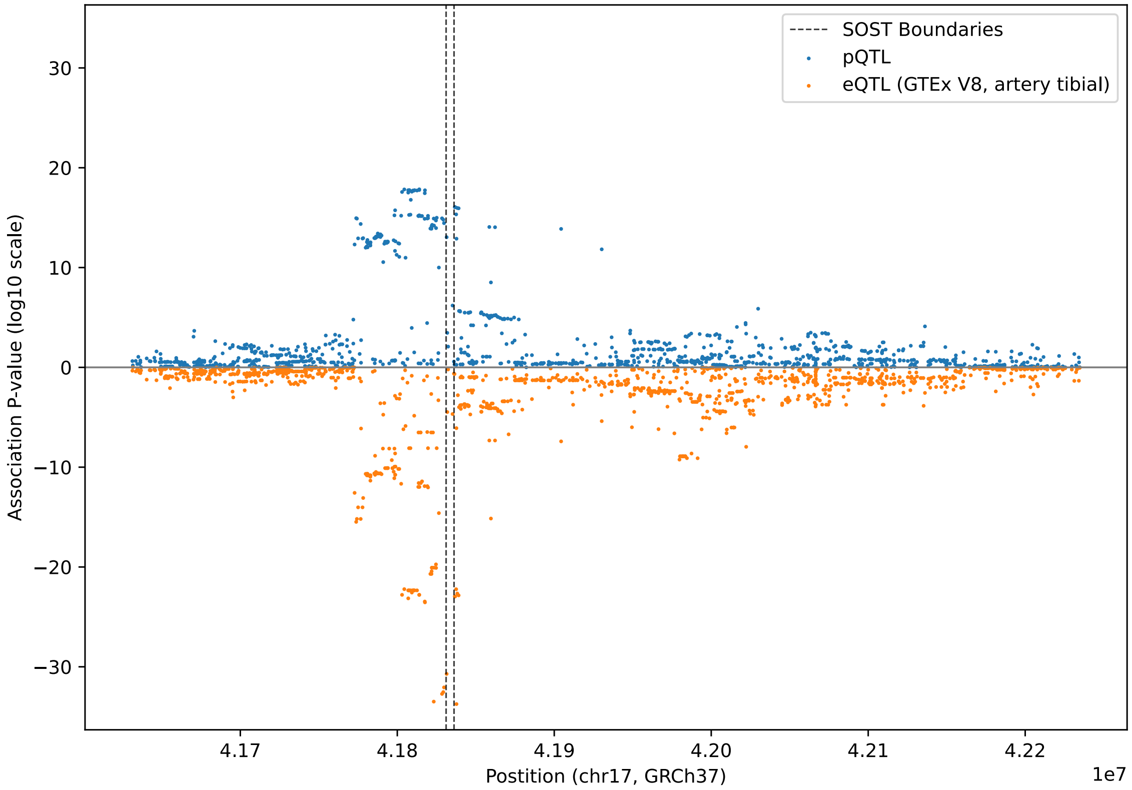


**Supplementary Figure A7: Association P-values (log scale) between genetic variants at the *SOST* locus and *SOST* expression in the tibial artery in GTEx V8 and circulating sclerostin levels in the UK Biobank.**

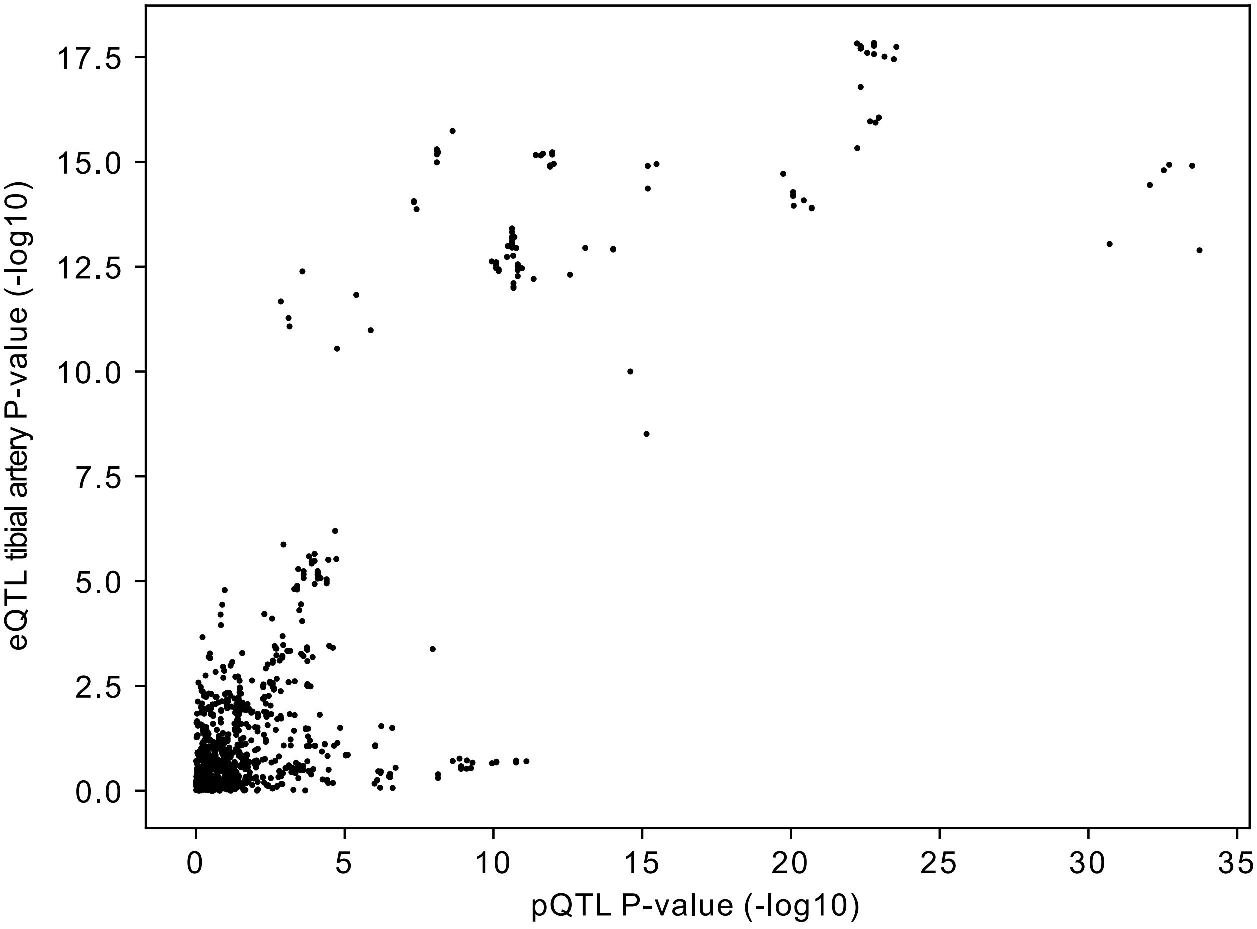


**Supplementary Figure A8: Comparison of sclerostin tibial artery eQTL and pQTL P-values (log scale).**

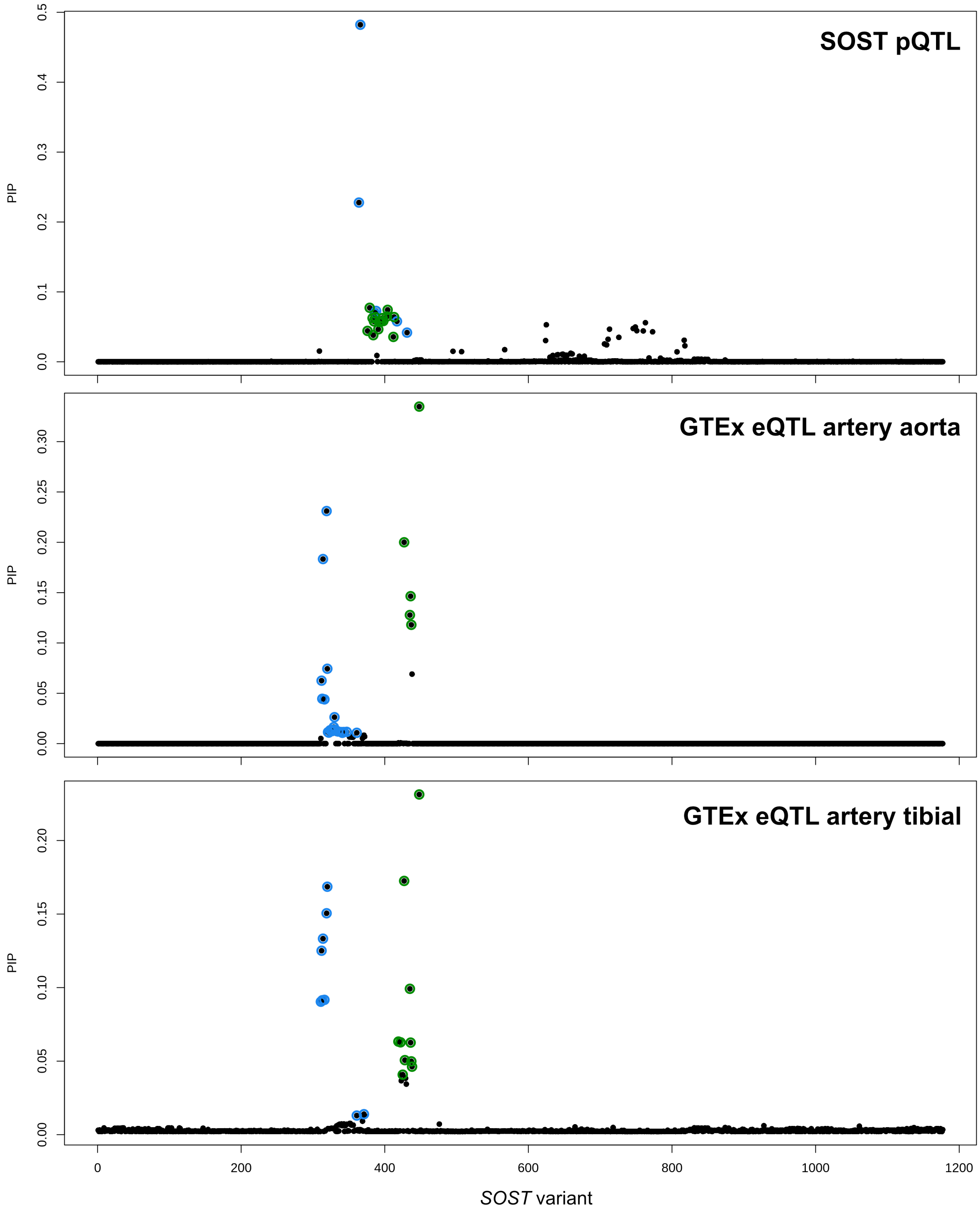


**Supplementary Figure A9: Posterior inclusions probabilities of variants at the *SOST* locus in finemapping credible sets inferred by SuSiE for association with circulating protein levels (UK Biobank) and sclerostin expression (aorta and tibial artery, GTEx).**

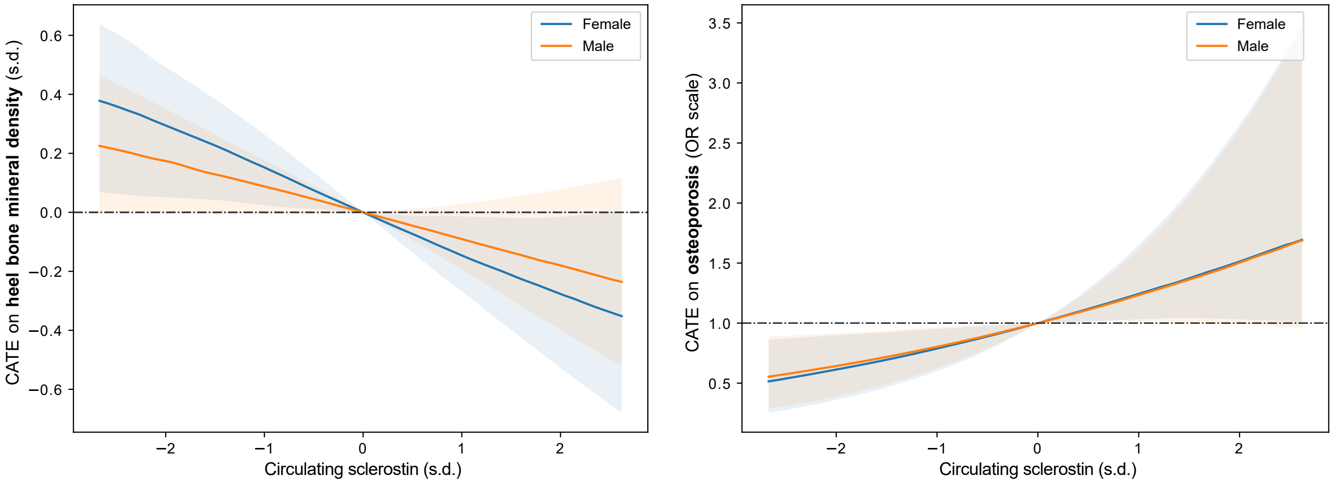


**Supplementary Figure A10: Conditional average treatment effect of circulating sclerostin levels on heel bone mineral density and osteoporosis in men and women estimated using Quantile IV in the UK Biobank.** The shaded region corresponds to 90% bootstrap confidence intervals. The plots cover the central 99% of the exposure range.

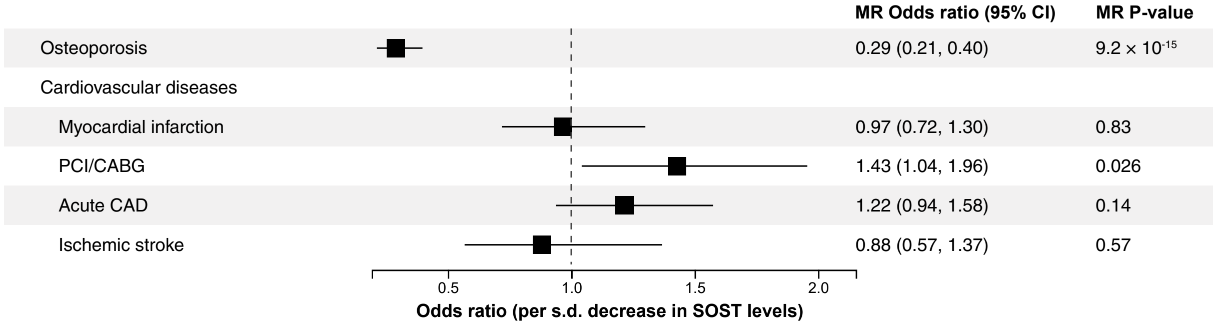


**Supplementary Figure A11: Mendelian randomization of the effect of a 1 s.d. reduction in circulating sclerostin using the Inverse Variance Weighted estimator based on the finemapped sclerostin pQTLs in the UK Biobank**

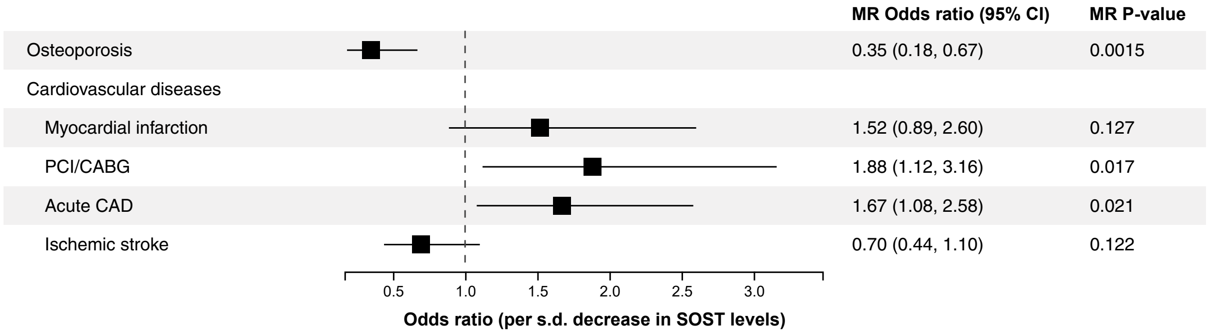


**Supplementary Figure A12: Mendelian randomization of the effect of a 1 s.d. reduction in circulating sclerostin using the PC-GMM estimator based on LD pruned variants at the *SOST* locus.**

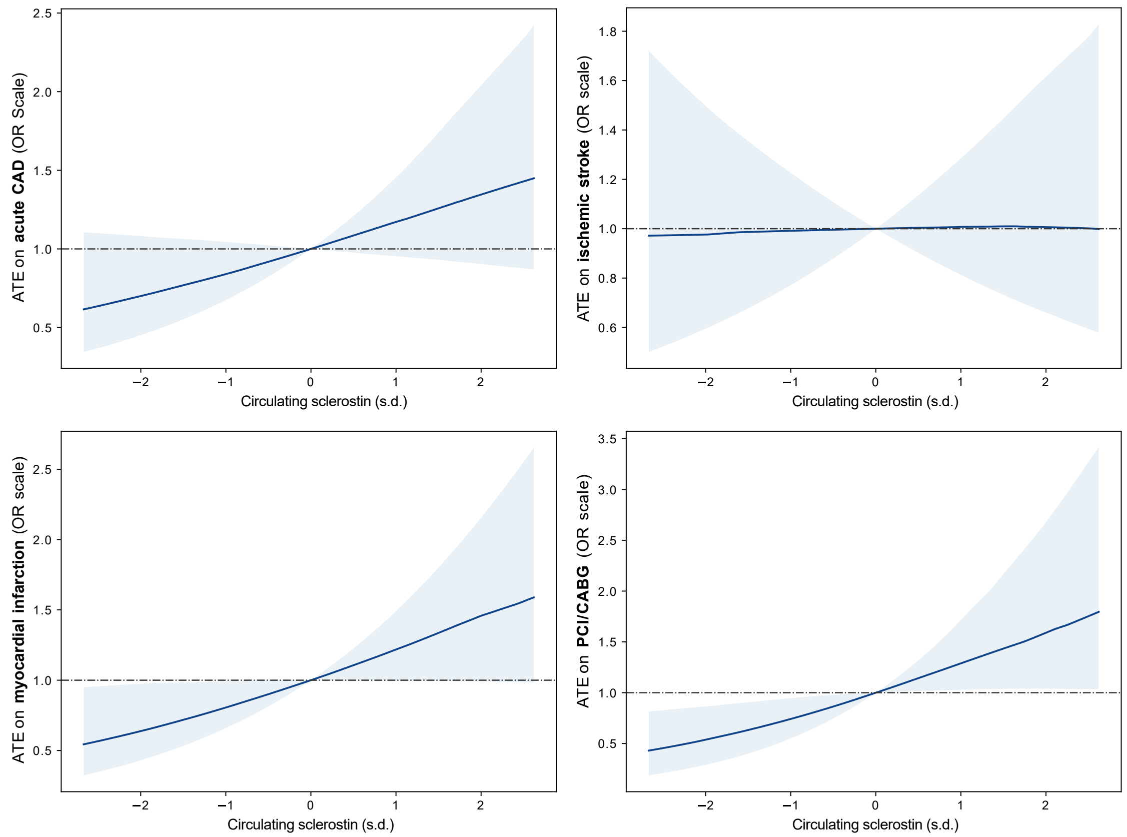


**Supplementary Figure A13: Quantile IV estimate of the average effect of varying the levels of circulating sclerostin about the mean on cardiovascular outcomes in the UK Biobank.** The shaded region corresponds to 90% bootstrap confidence intervals. The plots cover the central 99% of the exposure range.

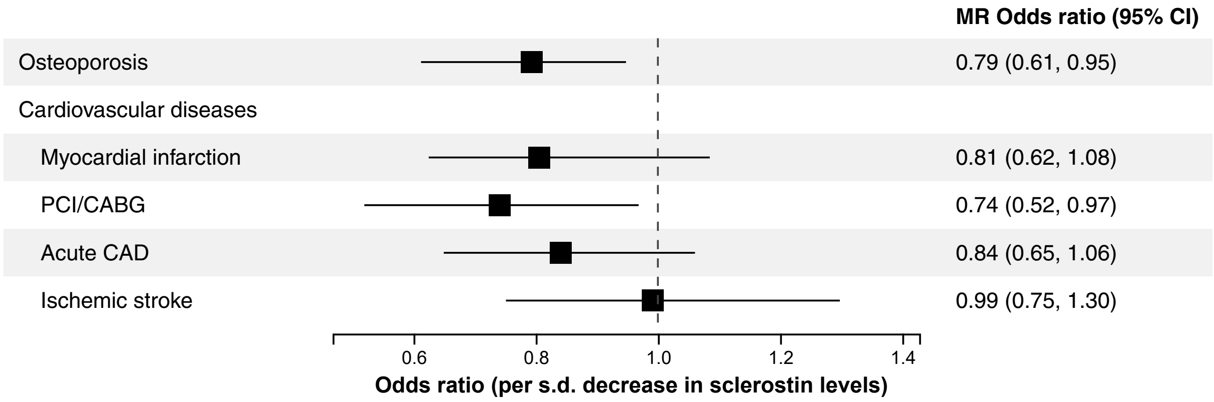


**Supplementary Figure A14: Mendelian randomization of the effect of a 1 s.d. reduction in circulating sclerostin using the Quantile IV estimator in the UK Biobank.**

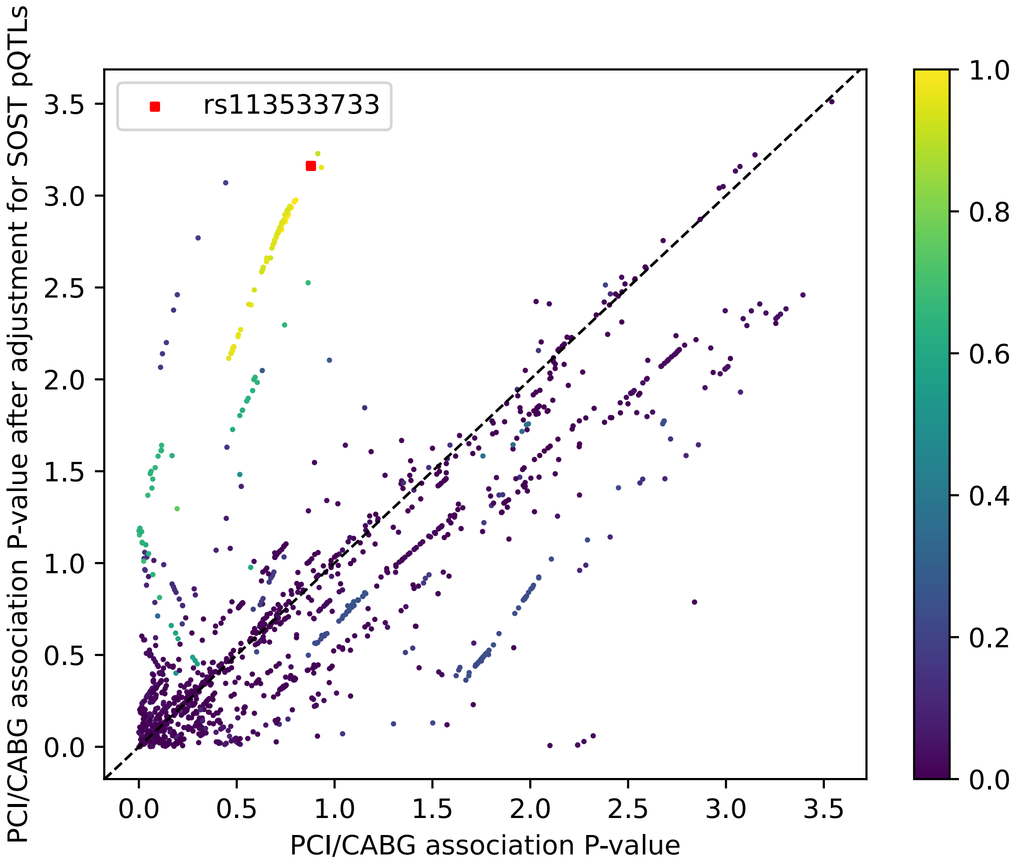


**Supplementary Figure A15: Comparison of the genetic association P-values before and after further adjustment for the sclerostin pQTLs.** The square on the scatter plot denotes the variant rs113533733 which we identified as having possible direct effects do to its high association P-value despite robust adjustment for variants associated with circulating sclerostin levels. The color represents LD ($r^2$) with this variant.

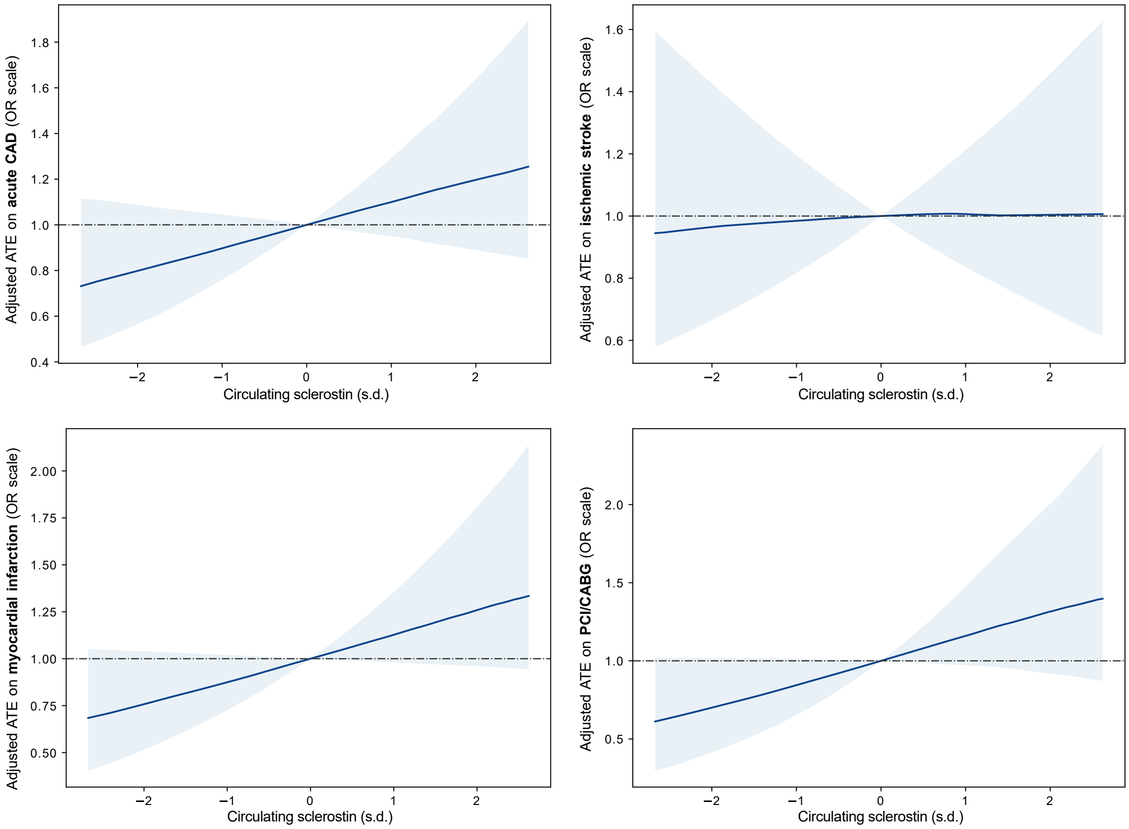


**Supplementary Figure A16: Adjusted Quantile IV estimate of the average effect of varying the levels of circulating sclerostin about the mean on cardiovascular outcomes accounting for direct effects by rs113533733 in the UK Biobank.** The shaded region corresponds to 90% bootstrap confidence intervals. The plots cover the central 99% of the exposure range.

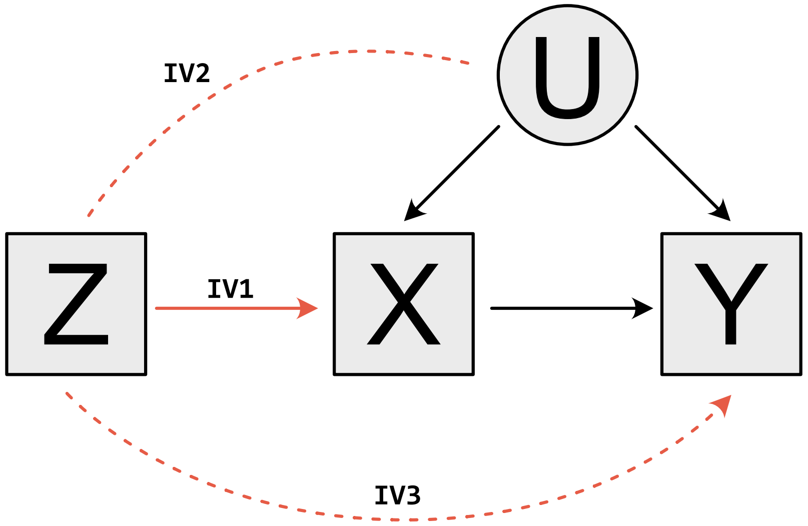


**Supplementary Figure A17: Graph illustrating the instrumental variable assumptions.** Dashed lines represent effects that are assumed absent. Solid lines denote effects that are assumed present. Squares denote observed variables and circles denote unobserved variables. When there are no arrow heads, the relationship is assumed to be bi-directional.
